# Brain Age Gap as Predictor of Disease Progression in Parkinson’s Disease

**DOI:** 10.1101/2025.10.15.25338068

**Authors:** Tom Hähnel, Shammi More, Felix Hoffstaedter, Kaustubh R. Patil, Holger Fröhlich, Björn Falkenburger

**Author notes:** <u>Corresponding author:</u> Dr. med. Tom Hähnel, Department of Neurology, Medical Faculty and University Hospital Carl Gustav Carus, TUD Dresden University of Technology, Fetscherstrasse 74, 01307, Dresden, Germany. Both authors contributed equally (Holger Fröhlich, Björn Falkenburger).

## Abstract

Parkinson’s disease (PD) exhibits high heterogeneity in disease progression, complicating management and increasing required sample sizes for clinical trials. This study evaluates Brain Age Gap (BAG)— the difference between brain age and chronological age—for predicting disease progression in PD. Structural MRI-derived gray matter volumes of 451 early disease stage PD patients and 172 healthy controls were analyzed. PD patients had a mean BAG of 1.1 years at baseline with fast-progressing patients exhibiting a BAG of 3.0 years, whereas slow-progressing patients resembled BAG of healthy controls. Higher BAG was associated with more severe baseline symptoms, faster cognitive decline in several domains, increased hazard of developing mild cognitive impairment, and faster progression of dopaminergic neuron loss in longitudinal DaTSCANs. BAG-based patient stratification could reduce sample sizes of randomized clinical trials by 26%–56%. These findings suggest BAG as a prognostic biomarker of disease progression, which may accelerate development of disease-modifying treatments.

## Introduction

Parkinson’s disease (PD) exhibits high heterogeneity in disease progression, posing challenges for clinical management and limiting the statistical power of clinical trials that investigate new, potentially disease-modifying drugs.^1^ PD heterogeneity has been explained by distinct progression subtypes, with individuals within each subtype showing more homogeneous progression patterns.^2–6^

Recently, a randomized clinical trial (RCT) investigating the effect of prasinezumab, a monoclonal antibody developed as a disease-modifying drug to slow PD progression, failed to reach its primary outcome.^7^ A secondary analysis revealed that the RCT could have demonstrated a significant treatment effect if patient inclusion had been restricted to a rapid-progressing PD subtype, which would have resulted in more homogeneous cohort and, on average, faster disease progression.^8^ Applied to multiple PD cohorts, similar stratification strategies have consistently improved statistical power.^3^ However, these approaches required longitudinal follow up prior study enrollment to generate reliable subtype predictions, limiting its feasibility.

Brain aging follows a temporal trajectory of structural changes that are reflected by brain atrophy patterns. Brain age can be estimated from structural magnetic resonance imaging (MRI) using gray matter volumes (GMV) or white matter volumes (WMV), but GMV has the most significant effect in brain age estimation.^9^ Brain age gap (BAG) is the difference between brain age estimated from structural MRI and chronological age. BAG has been suggested as a biomarker of overall brain integrity and is increased in neurodegenerative disorders.^9^ A positive BAG indicates that the estimated brain age exceeds the chronological age, i.e., that the brain structure resembles that of an older individual. Conversely, a negative BAG indicates that the brain structure resembles that of a younger individual. In people with Alzheimer’s disease (AD), BAG is increased by 3 to 8 years.^10^ Furthermore, BAG in AD predicts disease progression and conversion from mild cognitive impairment to dementia.^11–13^ Studies in PD have reported smaller BAG values, ranging from 0.7 years to 4.4 years.^14–24^ The potential for predicting disease progression and patient stratification in RCTs using BAG has not been explored.

We recently identified two PD progression subtypes using a data-driven approach and longitudinal data: a fast-progressing and a slow-progressing subtype.^3^ The fast-progressing subtype exhibited a more rapid progression in several cognitive domains, non-motor symptoms, specific gait patterns, and axial symptoms. Moreover, it was associated with worse response to dopaminergic treatment and increased mortality. In contrast, the slow-progressing subtype exhibited mild disease progression across most symptom domains and a more asymmetrical DaTSCAN uptake ratio. Notably, the existence of both subtypes was validated in multiple independent cohorts. However, longitudinal data was required for subtype predictions.

In this study, we aimed to assess the association of baseline BAG with disease progression in PD, using the two PD progression subtypes that were identified within the same dataset. First, we compared the atrophy patterns between the fast-progressing and slow-progressing PD subtypes directly using GMV derived from structural T1-weighted (T1w) MRI scans. To estimate BAG in our cohort, we applied multiple brain age estimation workflows from the literature and selected the one that provided the highest accuracy for our dataset. BAG estimates were then compared between PD progression subtypes and correlated with clinical progression rates, features extracted from DaTSCANs, and biomarkers of Alzheimer’s co-pathology. Finally, we analyzed if baseline BAG can be used to stratify patients and optimize clinical trial designs leading to reduced sample sizes. To minimize the risk of bias due to people with PD being at different disease stages, we corrected all analyses using a latent disease time approach published before ^3,25^.

## Results

### Demographics and clinical information

Overall, 451 people with PD and 172 healthy controls (HC) from the Parkinson’s Progression Markers Initiative (PPMI) cohort were included in our analysis (Table 1). HC and people with PD exhibited similar age and sex distributions. People with PD were at early disease stages as indicated by Hoehn & Yahr (H&Y) stages I and II observed in 443 of 451 of people with PD, low MDS-Unified Parkinson’s Disease Rating Scale (MDS-UPDRS) III scores (median: 20 points), and high Montreal Cognitive Assessment (MoCA) scores (median: 27 points). A minority of HC (39.0%, n=67) and people with PD (37.7%, n=170) had longitudinal MRI data available, whereas most participants had only a single baseline MRI. PD progression subtype assignments for people with PD were available from a previous analysis.^3^ Most people with PD were classified as slow-progressing (65.4%, n=295) and a smaller fraction as fast-progressing (14.6%, n=66) subtype. Because subtype identification required a longitudinal follow-up of specific clinical scores, no subtype information was available for a minority of people with PD (20.0%, n=90).

**Table 1:** Demographics and baseline clinical characteristics. Demographic and clinical characteristics of healthy controls and people with PD. Values are reported as median [interquartile range] or as relative frequency [absolute frequency]. Progression subtype assignments were available for 361 of 451 people with PD. Abbreviations: H&Y: Hoehn & Yahr, MDS-UPDRS: MDS-Unified Parkinson’s Disease Rating Scale, MoCA: Montreal Cognitive Assessment, SCOPA: Scales for Outcomes in Parkinson’s disease.

|  | Healthy controls<br>(n=172) | Parkinson's disease<br>(n=451) |
| --- | --- | --- |
| **Age (years)** | 61.4 [54.9-68.0] | 62.8 [55.3-69.1] |
| **Sex** |  |  |
| Male | 64.5% [111] | 62.5% [282] |
| Female | 35.5% [61] | 37.5% [169] |
| **H&Y** |  |  |
| H&Y 1 |  | 38.6% [174] |
| H&Y 2 |  | 59.6% [269] |
| H&Y 3 |  | 1.8% [8] |
| H&Y 4 |  | 0.0% [0] |
| H&Y 5 |  | 0.0% [0] |
| **Disease duration (years)** |  | 0.4 [0.2-1.0] |
| **Number of visits** | 14 [5-19] | 6 [4-14] |
| **Follow up (years)** | 5.0 [1.5-11.0] | 2.0 [0.9-7.0] |
| **MDS-UPDRS I** |  | 5 [3-8] |
| **MDS-UPDRS II** |  | 5 [3-8] |
| **MDS-UPDRS III** |  | 20 [15-26] |
| **MoCA** |  | 27 [26-29] |
| **SCOPA** |  | 12 [7-20] |
| **Number of MRI per individual** |  |  |
| Baseline only | 61.0% [105] | 62.3% [281] |
| 2 MRI | 27.9% [48] | 4.9% [22] |
| >2 MRI | 11.0% [19] | 32.8% [148] |
| **Subtype** |  |  |
| Fast |  | 14.6% [66] |
| Slow |  | 65.4% [295] |
| **Latent disease time (years)** |  | 0.2 [-0.9-1.2] |

### Correcting for different disease stages

A previously published latent disease time approach was used to correct subsequent analyses for differences in disease stages between people with PD.^3,25^ Therefore, we aligned people with PD on a latent disease timescale by comparing the individual progression of multiple motor and non-motor symptoms with the cohort’s average, yielding an estimated latent disease time for each person with PD that reflects the actual disease stage. Thereby, a latent disease time of 0 corresponds to the cohorts’ average disease stage at the time of diagnosis. Accordingly, positive or negative latent disease time values correspond to a disease stage that is later or earlier than the average disease stage at the time of diagnosis.

Applying this method, we estimated a mean latent disease time of 0.2 years at baseline visit, with an interquartile range between −0.9 and 1.2 years. (Table 1). Estimated latent disease times exhibited a stronger correlation with Hoehn & Yahr (H&Y) stages (τ=0.23, p<0.0001) compared to the correlation of time since diagnosis with H&Y (τ=0.12, p=0.0027), and this difference in correlations was significant (p=0.004), confirming the validity of our latent disease time approach.

### Voxelwise and parcelwise gray matter volume comparison of PD progression subtypes

First, we assessed whether a direct comparison of gray matter volumes (GMV) derived from structural MRI images at baseline can differentiate between fast-progressing and slow-progressing people with PD. GMV in voxelwise space, correcting for sex, age, total intracranial volume, and latent disease time, resulted in a visual pattern with a tendency towards more pronounced cortical atrophy for the fast-progressing subtype and more pronounced subcortical atrophy for the slow-progressing subtype (Fig. 1) However, no voxels were significantly different between the two progression subtypes after correction for multiple testing.

**Figure 1:**
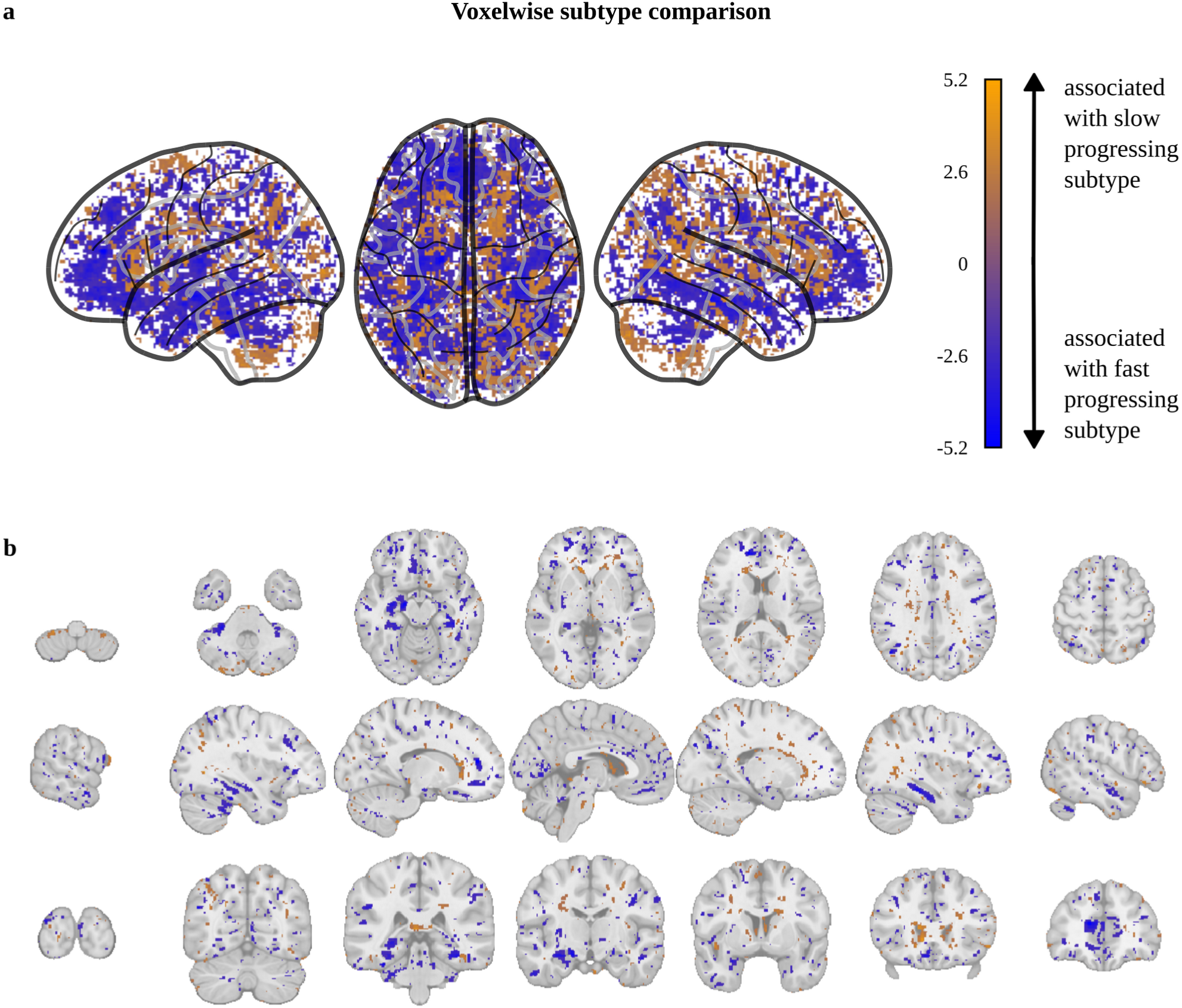
Voxelwise gray matter volume comparison of PD progression subtypes. Association of atrophy with the fast-progressing (blue) and slow-progressing (orange) subtypes displayed as **A)** glass brain visualization and **B)** sagittal, coronal, and transverse planes. Voxels with significant PD progression subtype differences before correction for multiple testing are colored depending on their t-value (blue: associated with fast-progressing subtype, orange: associated with slow-progressing subtype). Note that no voxels remained significant after Benjamini-Hochberg correction for multiple testing. No additional smoothing or resampling was performed for this analysis.

In a subsequent approach, we applied additional 4 mm resampling and 4 mm smoothing to voxelwise gray matter images to reduce noise and the number of statistical tests. This approach produced a similar visual pattern with no voxels reaching statistical significance between the two PD progression subtypes after correcting for multiple testing (Fig. S1). We then performed comparisons between PD progression subtypes using GMV in parcelwise space to further reduce the number of statistical tests. Again, no region exhibited significant differences after correcting for multiple testing, although several cortical regions showed a visual pattern towards more atrophy in the fast-progressing subtype (Fig. S2-5).

Overall, direct GMV comparison of structural MRI revealed visual patterns associated with PD progression subtypes but did not yield statistically significant differences.

### Brain age model selection and validation in HC

Motivated by these visual GMV patterns, we investigated whether brain age estimations using GMV as features at baseline could capture these patterns and provide information about clinical disease progression in PD.

Although several brain age models have been published, their accuracy varies depending on the datasets on which they were trained and the dataset on which they are applied.^26^ Moreover, brain age is typically overestimated in elderly individuals and underestimated in younger individuals. To remove this age-related bias, several bias correction methods have been proposed.^27^

Therefore, we systematically evaluated three brain age models from a recent systematic comparison in combination with two different bias correction methods in our dataset. Once the brain age was estimated by applying the three brain age models, bias correction was performed in cross-validation fashion in HC, where a BAG of zero is expected, to prevent overfitting (Table S1). The brain age model using lasso regression with no smoothing and 4-voxel resampling (S0_R4+LR), combined with bias correction via BAG and chronological age (Beheshti method^27^) achieved the best accuracy. This workflow yielded a mean absolute error of 4.51 years (Fig. S6), consistent with the accuracy of brain age models in the literature.^26,28^ Therefore, the S0_R4+LR brain age model was used for all further analysis, with the Beheshti bias correction method being retrained on all HC.

Applying this workflow to the complete HC group, brain age correlated strongly with chronological age (ρ=0.90, p<0.0001), with a mean absolute error of 4.34 years (Fig. 2A). As expected, there was no significant difference between mean brain age and mean chronological age in HC (BAG=0.00 years, p=1, Fig. 2B) and BAG was not correlated with chronological age (ρ=0.00, p=1, Fig. S6), indicating a successful bias correction. The concordance correlation coefficient for BAG in participants with short-term follow-up MRI was 0.86, demonstrating good re-test reliability of our brain age estimations. Longitudinal consistency of brain age was confirmed by a significant correlation between the increase in brain age between two subsequent MRI scans with the actual time elapsed between them (ρ=0.51, p=0.016, Fig. 2C).

**Figure 2:**
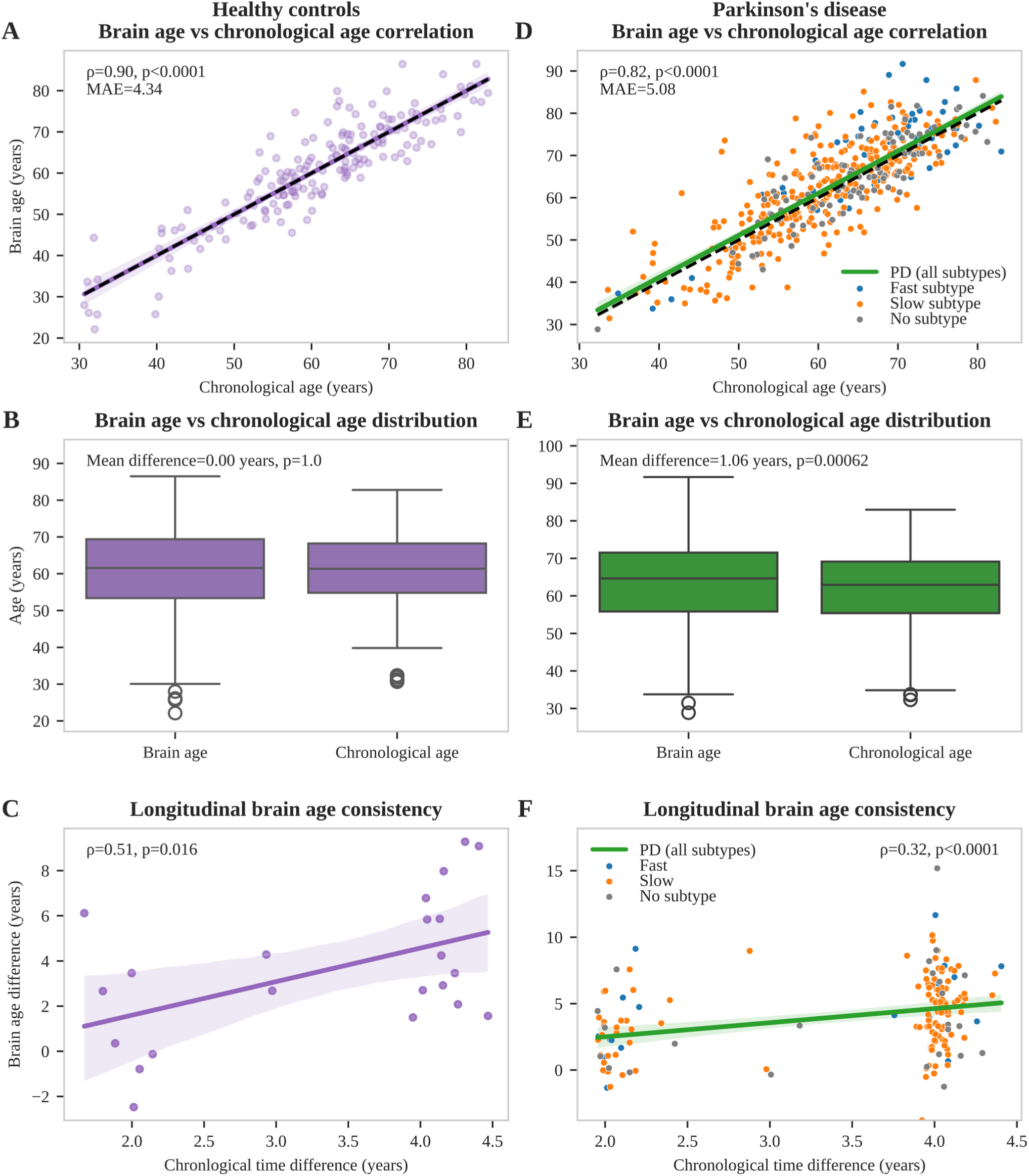
Brain age calculation in healthy controls and Parkinson’s disease. Brain age estimations for healthy controls (purple) are shown in the left column, brain age estimations for the PD cohort (green) in the right column. Individual people with PD are colored depending on their progression subtype (blue: fast-progressing, orange: slow-progressing, gray: no subtype available). **A/D:** Correlations of brain age with chronological age. Mean absolute error, Pearson correlation coefficient and corresponding p-value are shown. **B/E:** Box plots comparing distributions of chronological age and brain age. P-values of corresponding paired t-tests are shown together with the mean age differences. **C/F:** Longitudinal consistency analyses showing correlations of brain age difference and chronological age difference between subsequent MRIs. Pearson correlation coefficient and corresponding p-value are reported. All brain age estimations were based on a bias correction trained on the whole HC group to allow comparison between HC and PD. Cross-validation results for HC are presented in Fig. S6 and were used for model selection. Abbreviations: PD: Parkinson’s disease, MAE: mean absolute error.

Overall, the selected brain age estimation workflow demonstrated state-of-the-art performance compared to recent literature.^26,28^

### Brain age estimation in PD

When applied to people with PD, brain age estimations correlated strongly with chronological age (ρ=0.82, p<0.0001), with a mean absolute error of 5.08 years (Fig. 2D), slightly higher than that observed in the HC group. Estimated brain ages were significantly higher than chronological ages (p=0.00062, Fig. 2E), resulting in a mean BAG of 1.06 years. As in HC, BAG did not correlate with chronological age (ρ=-0.01, p=0.91), indicating successful bias correction also in PD. The concordance correlation coefficient was 0.89, indicating good re-test reliability of BAG estimations. Longitudinal consistency of brain age estimations was confirmed by a significant correlation between the increase in brain age between two subsequent MRI scans with the actual time elapsed between them (ρ=0.32, p<0.0001, Fig. 2F).

Overall, the selected brain age estimation workflow provided reliable BAG estimations, with a mean BAG of 1.06 years in PD being consistent with literature reports.^14–20^

### Brain age gap in fast-progressing and slow-progressing PD

We then assessed whether BAG serves as a marker of disease progression by comparing BAG between PD progression subtypes.

The fast-progressing PD subtype exhibited a significantly higher BAG at baseline than HC (ΔBAG=3.00 years, p=0.0009, Fig. 3A) and the slow-progressing PD subtype (ΔBAG=2.02 years, p=0.045). No significant difference was observed between the slow-progressing and fast-progressing PD subtypes (p=0.11).

**Figure 3:**
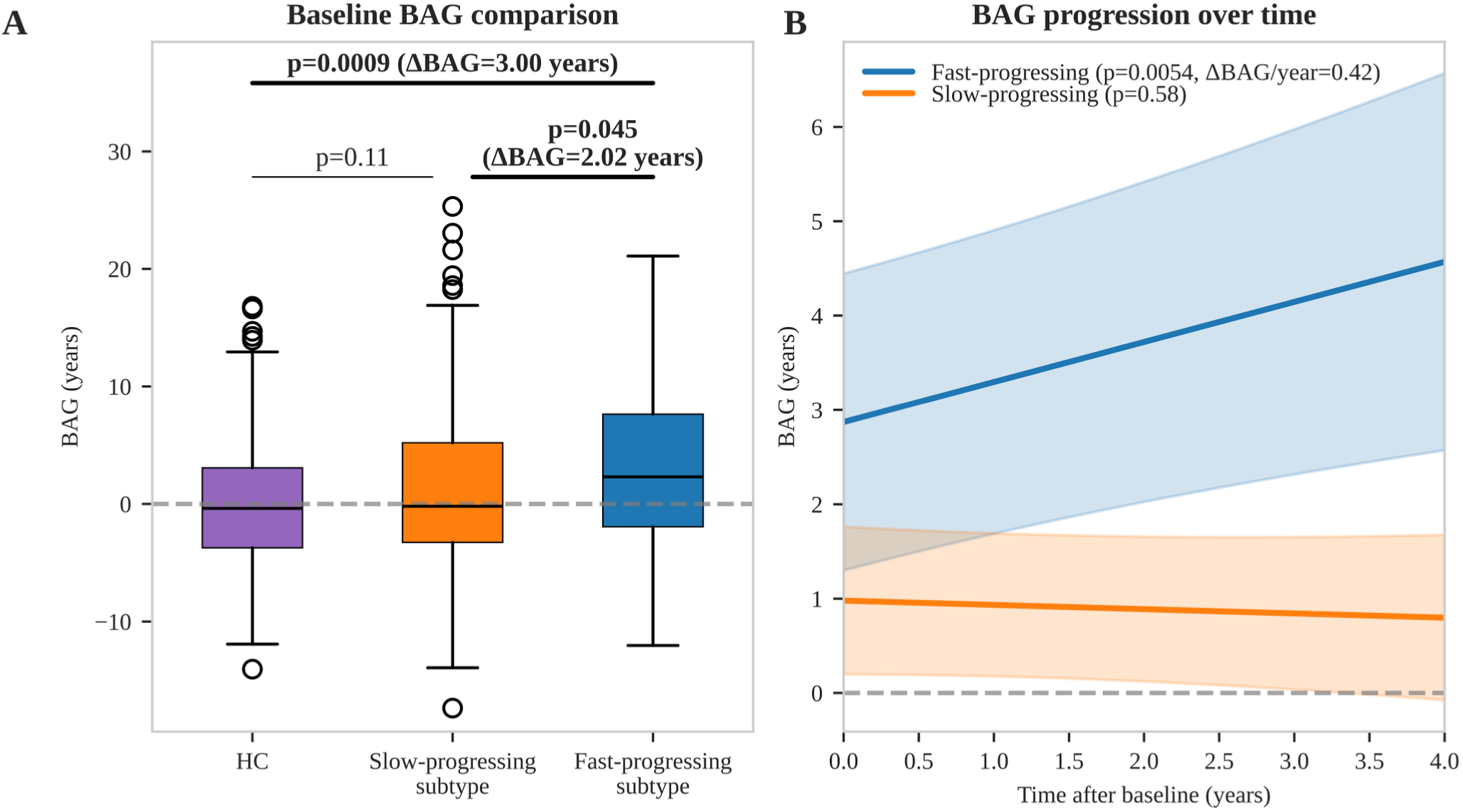
Brain age gap differences between PD progression subtypes and HC. **A:** BAG comparison between PD progression subtypes and HC at baseline. P-values for group differences, corrected for age and sex, are reported alongside the BAG difference for significant differences. The BAG difference between the slow and fast-progressing subtype was corrected for latent disease time as well. **B:** Longitudinal BAG increase estimated using a linear mixed effects model for each PD progression subtype. P-values for BAG progression over time are shown together with the mean BAG increase per year for significant results. Shaded areas indicate 95% confidence intervals. The horizontal dashed lines correspond to BAG = 0. Abbreviations: BAG: brain age gap. HC: healthy controls.

Longitudinal analysis of BAG trajectories revealed a significant annual increase of BAG in the fast-progressing PD subtype of 0.42 years per year (p=0.0054, Fig. 3B), meaning a 42% faster brain aging as it would be expected in HC. Conversely, the slow-progressing PD subtype showed no significant change in BAG over time (p=0.58).

### Brain age gap correlation with clinical scores

We now sought to distinguish for which clinical domains BAG represents a marker of disease severity and for which clinical domains a predictor of disease progression. Therefore, we evaluated correlations of baseline BAG with (I) baseline outcomes (reflecting disease severity) and (II) longitudinal progression of outcomes (reflecting disease progression).

Baseline BAG was correlated with cognitive scores, motor scores, and scores capturing overall disease severity (Fig. 4). Interestingly, BAG correlated with motor scores and overall disease severity scores at baseline while there was no association of BAG with progression of these scores. Conversely, BAG showed more significant correlations with cognitive decline over time than with baseline cognitive scores.

**Figure 4:**
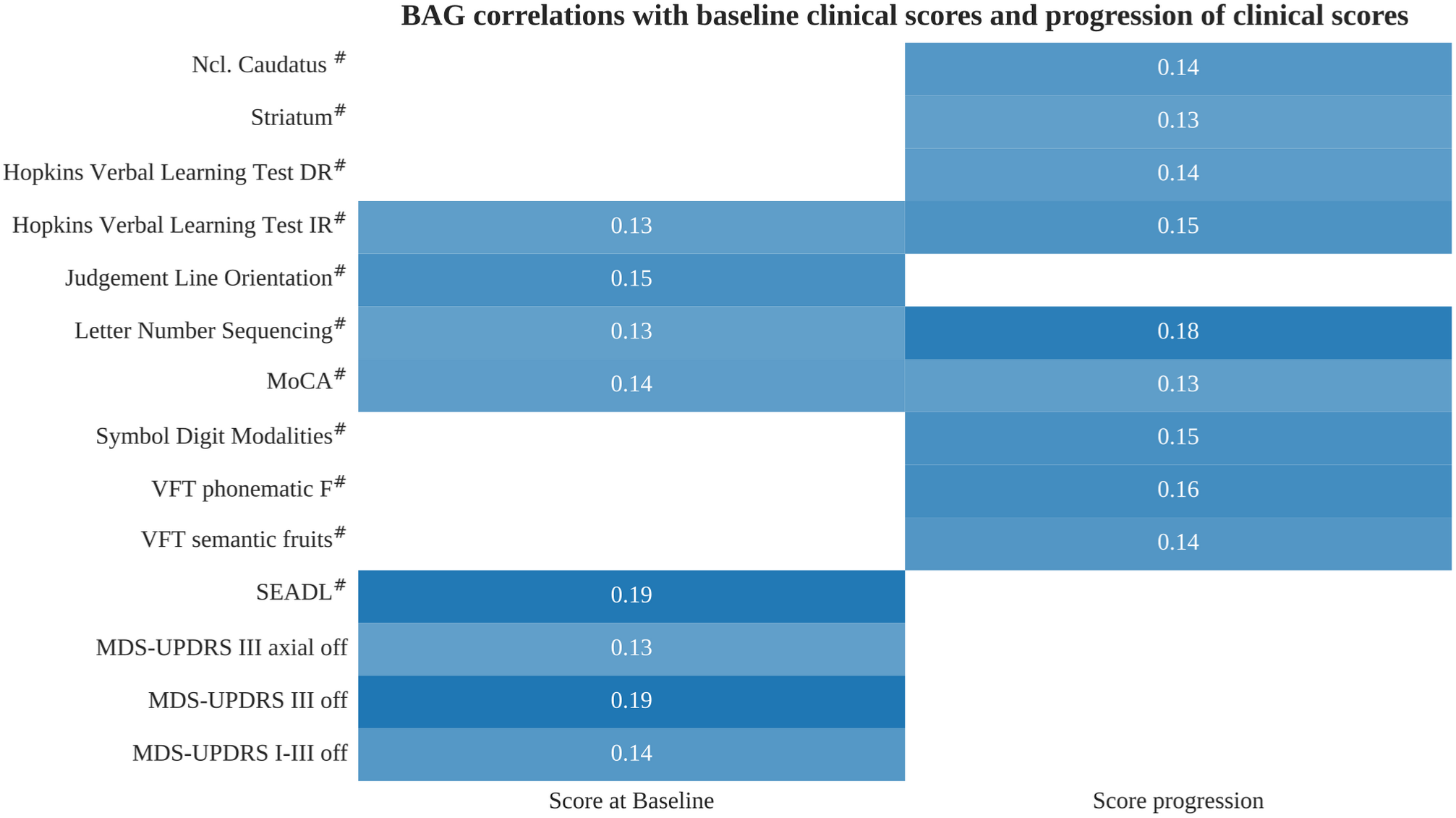
Brain age gap correlations with clinical scores. Correlations of BAG with baseline outcomes (left column) and progression of outcomes (right column) are depicted with the color indicating the strength of the partial correlation (light blue: low correlation, dark blue: high correlation). Partial correlation coefficients are shown and were corrected for age. Correlations with baseline outcomes were corrected for latent disease time as well. Only correlations with statistical significance after correction for multiple testing using Benjamini-Hochberg method are shown. Outcomes marked with # were inverted such that positive correlation coefficients always indicate higher BAG being associated with greater baseline severity or faster progression. A complete list, including also outcomes without significant correlations, is provided in Table S2-S4. Abbreviations: BAG: brain age gap, DR: delayed recall, IR: intermediate recall, MDS-UPDRS: MDS-Unified Parkinson’s Disease Rating Scale, MoCA: Montreal Cognitive Assessment, SEADL: Schwab and England Activities of Daily Living scale, VFT: Verbal Fluency Task.

No correlation of baseline BAG was evident for clinical scores reporting autonomic dysfunction, non-motor symptoms, anxiety, depression, sleep disturbances, and motor impairment in daily life (Table S2).

Analyzing baseline BAG correlations with biomarkers, we found higher BAG being correlated with faster degeneration of dopaminergic axon terminals in the caudate nucleus and overall striatum, as observed in longitudinal DaTSCANs. Contrarily, BAG was not correlated with DaTSCAN uptake ratios in these regions at baseline (Table S3). Furthermore, no significant correlations of BAG were found regarding baseline values and longitudinal changes of cerebrospinal fluid markers of Alzheimer’s disease pathology and neurofilament light chain (Table S4).

Using a Cox proportional hazards model as a complementary time-to-event approach, we identified higher BAG as a significant predictor for overall cognitive decline and progression to mild cognitive impairment (Fig. 5). In detail, each year increase in baseline BAG was associated with 2% increased hazard for cognitive decline (p=0.04) and 4% increased hazard for mild cognitive impairment (p=0.032). Visual and statistical checks indicated no violation of the proportional hazards assumption (Fig. S7).

**Figure 5:**
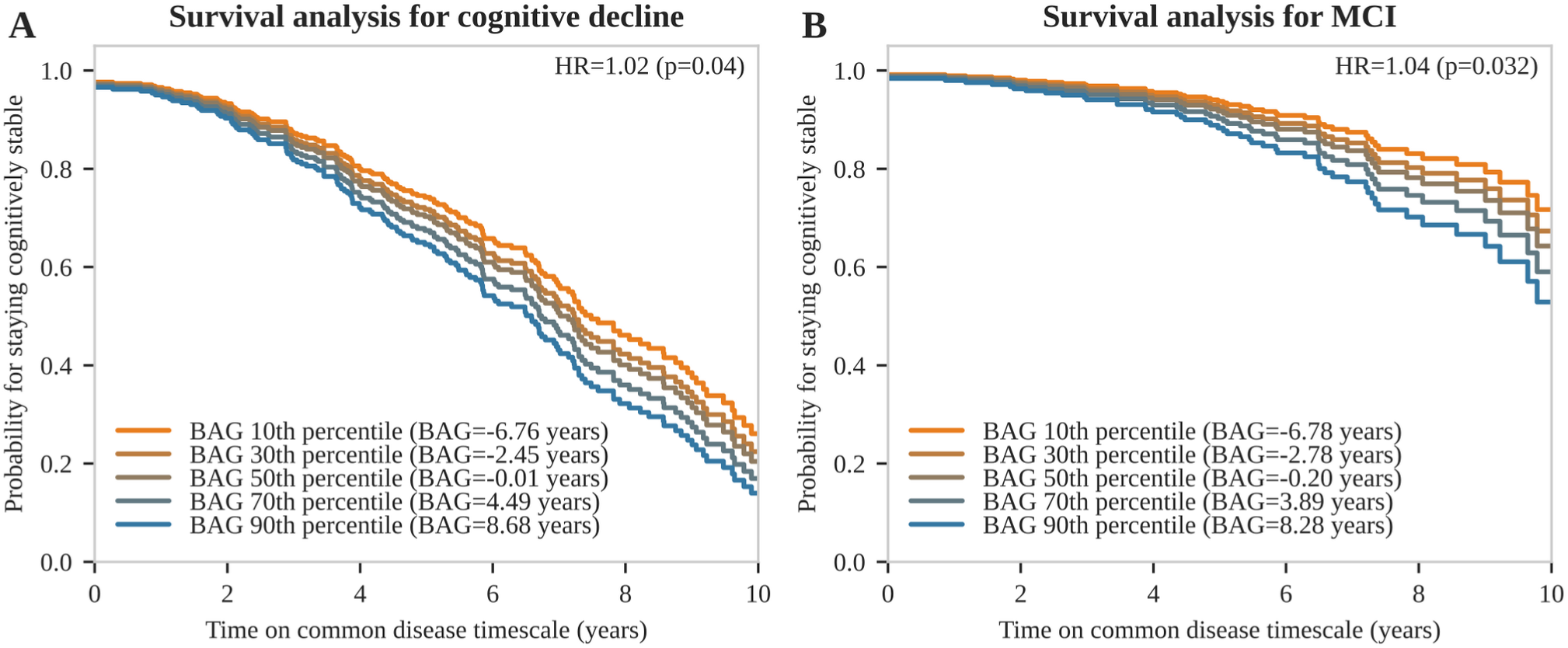
Event-free survival analysis for cognitive decline and mild cognitive impairment. Cox regression plots for **A)** cognitive decline and **B)** mild cognitive impairment depending on BAG percentiles at baseline and including age and sex as additional covariates. Hazard ratios per BAG increase of 1 year are shown with corresponding p-values. Cognitive decline was defined as a progression towards the next level on an ordinal scale containing the levels normal cognition, subjective cognitive impairment, mild cognitive impairment, and dementia. Abbreviations: BAG: brain age gap, HR: hazard ratio, MCI: mild cognitive impairment

Overall, these findings suggest that BAG is predictive of overall cognitive decline, decline in several cognitive domains, and faster neurodegeneration observed in longitudinal DaTSCANs.

### Clinical trial optimization using brain age gap

Motivated by the finding that BAG correlates with cognitive decline, we assessed how baseline BAG can be used to optimize clinical trial designs by patient stratification.

We simulated a randomized clinical trial (RCT) assessing the effect of a disease-modifying drug on cognitive decline in people with PD. We assumed a 30% treatment-induced reduction of cognitive decline, a two-year follow-up period, and visits every six months. Cognitive decline was measured using a cognitive compound score derived from four cognitive tests. Without stratification, i.e., including all people with PD, 1218 people with PD were required to achieve 80% statistical power (Fig. 6). Applying a BAG-based stratification–including only people with PD with a BAG above the 50^th^, 70^th^, and 90^th^ percentile–reduced the required sample sizes by 23% (n=930), 28% (n=874), and 58% (n=507), respectively.

**Figure 6:**
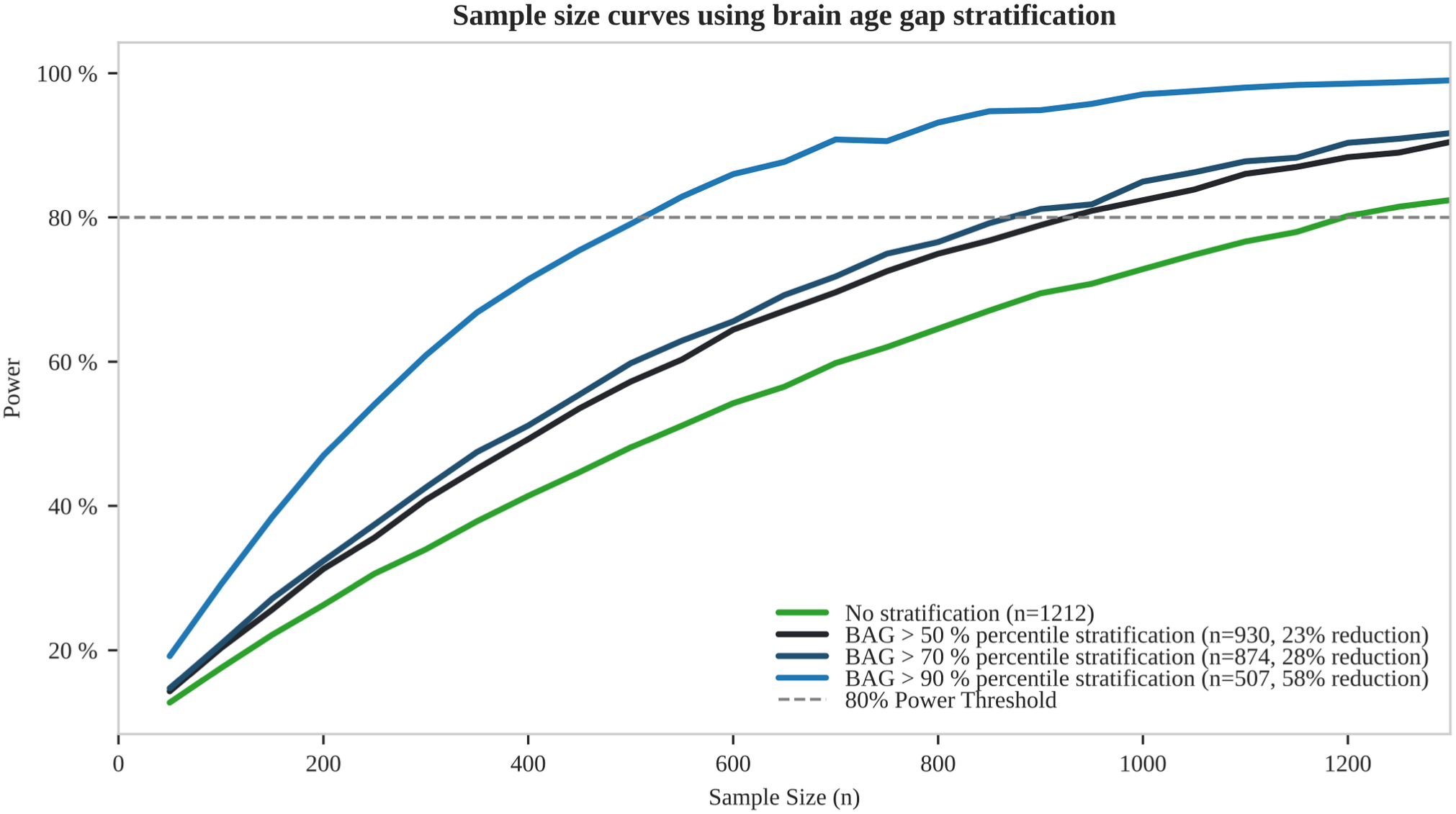
Sample sizes required for simulated clinical trials using brain age stratification. Estimated power and sample size curves for a clinical trial depending on the BAG threshold used as inclusion criterion: no stratification applied to people with PD (green), inclusion of people with PD with a baseline BAG greater than the 50^th^ percentile (black), 70^th^ percentile (dark blue), and 90^th^ percentile (light blue). A treatment effect of 30% on the progression rate of the cognitive composite score, a two-year observation period, and a significance level α=0.1 were assumed. Sample sizes and relative reductions compared to the unstratified scenario are reported for 80% power (dashed line). Abbreviations: BAG: brain age gap.

Repeating this analysis with single cognitive scores instead of the cognitive composite score, sample size reductions were found for all cognitive outcomes with median reductions being even higher: 44% (50^th^ BAG percentile), 54% (70^th^ BAG percentile), and 70% (90^th^ BAG percentile), indicating that our approach is not limited to a specific cognitive score. However, absolute sample sizes were significantly higher for single cognitive scores compared to the cognitive compound score (Table S5).

## Discussion

Our work demonstrates that GMV from structural MRI scans convey information about disease progression in PD. Traditional voxelwise or parcelwise GMV analysis using linear models and adjusting for important covariates cannot detect robust atrophy patterns. However BAG models provide a new, machine learning-based approach for capturing disease-related patterns in MRI scans. While a mean BAG of about 1 year in PD aligns with recent literature, our work provides additional insight that increased BAG is driven by a small fraction of fast-progressing people with PD, whereas BAG in most people with PD resembles that of HC. Additionally, BAG increased by 0.42 years per year in fast-progressing people with PD, leading to even larger differences between progression subtypes over time. Higher BAG was associated with more severe symptoms at baseline and predicted faster cognitive decline and faster degeneration of dopaminergic axon terminals from the substantia nigra. PD patients can be stratified based on their baseline BAG to enrich clinical trials with fast cognitive declining patients. Thereby, sample sizes of clinical trials could be reduced by 23%-58% in our simulations, depending on the fraction of patients that can still be included.

Our analysis showed a mean BAG difference of 1.06 years in PD. This value is within the lower range of BAG values reported for PD in the literature (0.7-4.4 years)^14–23,29,30^ and notably lower than BAG in Alzheimer’s disease (about 3-8 years).^10^ The relatively low BAG value compared to other PD publications may be due to PPMI inclusion criteria which restricted participants to the early disease stage with a maximum of 2 years since diagnosis. Our results and those of other authors indicate that BAG increases with disease duration for at least some patients, potentially leading to higher BAG values in advanced disease stages.^19,21,23^

Furthermore, the use of mean absolute error as model selection criterion has been the subject of controversy, given that complex machine learning methods used in brain age models are more likely to overfit the training data.^26,31^ Thus, these models may not generalize well to neurodegenerative disorders, contrary to providing lower BAG in the training cohort. As we (1) performed model selection on a dataset that was not used to train the brain age model and (2) selected the bias correction method using cross-validation, an overfit is relatively unlikely. Additionally, BAG in people with PD estimated by the other brain age workflows exhibited only minimal differences compared to the approach that provided the minimal mean absolute error (Table S6).

Our findings are in line with previous publications of BAG being associated with scores reporting motor symptoms^18,21,23,29,32^, cognitive impairment^15,18,21–23,29,32^, and overall disease severity^19,30^. However, we did not observe correlations between BAG and cerebrospinal fluid markers of Alzheimer’s dementia, neurofilament light chain, or uptake ratios extracted from DaTSCAN analysis at baseline, indicating that BAG provides an independent biomarker. In line with this, Teipel et al. found only minimal BAG differences in LRRK2 and GBA PD subtypes, indicating that BAG values may be also independent of genetic causes of PD.^18^

Literature on BAG as a predictor of disease progression in PD is sparse. It has been shown that BAG is higher in people with PD who converted to MCI than in people with PD who remained cognitively unimpaired.^22^ While this could have reflected the association of higher BAG being found in advanced disease stages, our study demonstrates that BAG is truly predictive of a faster rate of cognitive decline with the hazard of MCI being increased by 4% per each BAG year.

Higher BAG values at baseline have been reported as predictors of faster decline in visuo-executive functions (Symbol Digit Modalities Test), verbal learning and memory (Hopkins Verbal Learning Test), and overall cognition (Montreal Cognitive Assessment, MoCA).^18^ While our analysis confirms these findings, we provide additional evidence that high BAG values also predict faster cognitive decline in executive function/language (semantic and phonematic Verbal Fluency Tasks), and attention/concentration (Letter Number Sequencing test) domains. Thus, BAG should be interpreted as a predictor of overall cognitive decline in PD rather than being restricted to a specific cognitive domain.

Importantly, there was no association between Aβ42 or phospho-tau-levels and BAG, indicating that BAG reports cognitive decline independent of the co-occurrence of Alzheimer pathology. Moreover, BAG was not correlated with neurofilament light chain, indicating that brain atrophy patterns leading to increased BAG are not primarily caused by an overall faster neurodegeneration. Conversely, BAG at baseline was associated with faster decline of uptake ratios observed in longitudinal DaTSCAN measurements.

While higher BAG was correlated with faster progression of several cognitive scores, we found no correlation with progression of non-cognitive domains including motor symptoms and other non-motor symptoms. Therefore, BAG should be considered one of several factors contributing to the fast-progressing subtype, primarily explaining its cognitive characteristics.

We compared BAG between two PD progression subtypes that were validated in multiple external cohorts before.^3^ BAG was significantly elevated only in the fast-progressing subtype, which comprises a small fraction of people with PD (18%) and exhibited 42% faster brain aging than HC. Surprisingly, in the slow-progressing subtype, which represents the majority (82%) of people with PD, BAG was not elevated and did not increase over time. These findings support the notion that PD should be viewed as distinct subtypes with potentially different biological origins, rather than a continuum, and that BAG serves as a biomarker of the fast-progressing PD subtype.

Our voxelwise GMV comparison between PD progression subtypes indicated a visually pronounced cortical atrophy in the fast-progressing subtype and subcortical atrophy in the slow-progressing subtype. These visual atrophy pattern are in accordance with the higher cognitive impairment and faster cognitive decline observed in the fast-progressing PD subtype.^3^ A potential biological explanation behind these visual patterns may be provided by the α-synuclein Origin and Connectome Model (SOC Model) which expects that the body-first subtype exhibits already more severe and bilateral cortical α-synuclein pathology at the time of diagnosis than the brain-first subtype.^2,33^ Notably, the body-first subtype has been discussed as a potential biological explanation behind the fast-progressing subtype as it exhibits similar clinical characteristics.^3^ However, other hypothesis like distinct α-synuclein strain types have also been discussed as biological drivers of PD progression subtypes.^34^

The Federal Drug Agency (FDA) acknowledges prognostic enrichment strategies as they may increase the efficiency of drug development and support precision medicine (see “Enrichment Strategies for Clinical Trials to Support Approval of Human Drugs and Biological Products - Guidance for Industry”, www.fda.gov/media/121320/download, published in 2019). By enriching cohorts with patients who are likely to experience the outcome of interest (i.e., clinical progression), this strategy allows to reduce the sample size required for RCTs. Here, we showed that BAG-based prognostic enrichment lowers the required sample size in RCTs by 23-58%, while still allowing inclusion of 10-50% of people with PD. Our RCT simulations using cognitive primary endpoints are in line with the ongoing PreCoDe trial that investigates the effect of prasinezumab on a cognitive composite score in PD.^35^ However, most previous RCTs have used a motor or combined endpoint^7,36^.

Our presented enrichment strategy allows patient stratification based on a single and objective parameter obtained at baseline. This provides a complementary approach to clinical trial optimization strategies we previously published: (I) using longitudinal follow-up of multiple clinical scores to enrich RCTs with fast-progressing PD patients, and (II) using primary outcomes derived from digital biomarkers assessing motor functions.^3,37^ Further research should systematically investigate combinations of different clinical trial optimization strategies. Importantly, BAG-based enrichment strategies could be applied to other neurological diseases like Alzheimer’s disease where an association of BAG with disease progression has been observed.^12,13,38^

Our results are based on the PPMI cohort which restricts PD patients to the early disease stage at inclusion. While this demonstrates the high prognostic potential of BAG in early disease stages, it offers only limited insight into PD in advanced disease stages. For example, it remains unclear whether BAG remains consistently low in slow-progressing people with PD or whether it will increase in later disease stages. Thus, validation of our findings in cohorts with advanced stage PD is warranted.

We limited our systematic comparison of brain age models to workflows based on GMV. Recent studies have shown that brain age estimations might be slightly more accurate if a combination of GMV, WMV, and cerebrospinal fluid volumes is used.^26^ Furthermore, regional brain age models have been proposed, potentially providing better interpretability by restricting features to specific regions of interest.^16,39^

Although BAG shows promise for patient stratification in clinical trials, its utility for predicting outcomes in individual people with PD is limited, as most BAG estimations in our analysis fall within the model’s mean absolute error of 4.34 years. Integrating multimodal data has been shown to increase accuracy in predicting individual clinical outcomes in PD.^40^ Further research should therefore explore whether combining BAG and other data modalities (e.g., digital biomarkers) could improve predictions of disease progression at the individual level.

Our study demonstrates that BAG is a prognostic biomarker for faster neurodegeneration and cognitive decline in PD, particularly distinguishing fast-progressing people with PD. BAG offers potential to optimize clinical trial designs, thereby accelerating the development of new disease-modifying drugs.

## Methods

### PPMI cohort

We analyzed HC and people with PD from the Parkinson’s Progression Markers Initiative (PPMI, NCT04477785) with visits between 2010 and 2024. PPMI included HC and untreated, de-novo PD patients whose clinical diagnosis was confirmed by a pathological dopamine transporter SPECT (DaTSCAN). Furthermore, PD inclusion was limited to individuals with a clinical diagnosis within two years before the baseline visit. Written informed consent for data collection and sharing was obtained from all participants by PPMI. Ethical guidelines on human data collection were adhered to. The PPMI project was approved by the Institutional Review Board or Independent Ethics Committee of all participating sites in Europe, including Attikon University Hospital (Greece), Hospital Clinic de Barcelona and Hospital Universitario Donostia (Spain), Innsbruck University (Austria), Paracelsus-Elena-Klinik Kassel/University of Marburg (Germany), Imperial College London (UK), Pitié-Salpêtrière Hospital (France), University of Salerno (Italy), and in the USA, including Emory University, Johns Hopkins University, University of Alabama at Birmingham, PD and Movement Disorders Center of Boca Raton, Boston University, Northwestern University, University of Cincinnati, Cleveland Clinic Foundation, Baylor College of Medicine, Institute for Neurodegenerative Disorders, Columbia University Medical Center, Beth Israel Medical Center, University of Pennsylvania, Oregon Health and Science University, University of Rochester, University of California at San Diego, and University of California, San Francisco.

### Parkinson’s disease progression subtypes

PD progression subtypes were identified in a recent publication using a latent time joint mixed-effects model (LTJMM) and variational deep embedding with recurrence (VADER) in the PPMI cohort.^3^ PD subtype assignments were available for PD patients with longitudinal assessments of non-motor and motor scores (i.e., at least two visits with MDS-UPDRS I-IV, MoCA, PIGD score, and SCOPA-AUT).

### Latent disease time calculation

Disease progression was modeled as a multivariate linear process using a latent time joint mixed-effects model (LTJMM) as described in previous publications.^3,25^

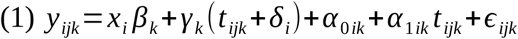

We denoted *y_ijk_* as outcome *k* measured at visit *j* for individual *i*. We adjusted for age at diagnosis and sex using covariates *x_i_* with coefficients *β_k_* shared across individuals. Cohort-level progression *γ_k_* was represented by a fixed mean slope for each outcome *k*, common to all individuals. Time was measured as time since diagnosis *t_ijk_*. To align trajectories across people people with PD, we introduced an individual-specific time shift *δ_i_* shared across outcomes, so trajectories were defined on a common latent disease time (*t_ijk_* + *δ_i_*). Individual heterogeneity was modeled with random intercepts *α_0ik_* and random slopes *α_1ik_*. Measurement errors *ε_ijk_* and time shifts *δ_i_* were assumed normal distributed with mean zero, and random effects (*α_0ik_*, *α_1ik_*) followed a multivariate normal distribution with mean of zero.

Models were fitted via Markov chain Monte Carlo (MCMC) with 4 chains, 25,000 iterations and 12,500 warm-up steps using the R packages ltjmm^41^ and rstan^42^. All outcomes were min–max normalized to their theoretical ranges; MoCA scores were inverted to ensure positive slopes across outcomes. Convergence was assessed visually and using *R̂* statistics, ensured to be below 1.05. The following outcomes were used in LTJMM to capture progression of a wide variety of motor and non-motor scores: MDS-UPDRS I-IV, Postural Instability and Gait Dysfunction score (PIGD), MoCA and Scales for Outcomes in Parkinson’s Disease-Autonomic Dysfunction (SCOPA).

To validate our LTJMM approach, we calculated Kendall tau-b rank correlation coefficients for disease stages reported by H&Y stages with (I) time since diagnosis and (II) latent disease times. The difference between the two correlations was assessed statistically using a bootstrapping approach.

### MRI acquisition

For training the brain age estimation model, four large neuroimaging datasets were used: Cambridge Centre for Ageing and Neuroscience (CamCAN, N = 651)^43^, Information eXtraction from Images (IXI, N = 562) (https://brain-development.org/ixi-dataset/), the enhanced Nathan Kline Institute-Rockland Sample (eNKI, N = 597)^44^, and the 1000 brains study (1000BRAINS; N = 1143)^45^ as described in a recent publication.^26^ MRI inclusion was restricted to HC between ages of 18 and 90 years with no diagnosis of psychiatric, neurological, or major medical conditions.

PPMI T1-weighted (T1w) MRIs were acquired for HC and people with PD using 3D volumetric sequence (e.g., MP-RAGE, IR-FSPGR) in the sagittal plane, with a slice thickness of 1.2 mm or less and details being reported online (https://www.ppmi-info.org/study-design/research-documents-and-sops).

### MRI preprocessing

T1w images were preprocessed using the Computational Anatomy Toolbox (CAT) 12.8^46^, replicating the workflow of a recent publication.^26^ For precise normalization and segmentation, T1w images first underwent affine registration with increased accuracy (accstr = 0.8). Following bias field correction and tissue class segmentation, normalization was performed using an optimized Geodesic Shooting approach with regstr = 1.^47^ We utilized 1 mm Geodesic Shooting templates and generated output images at 1 mm isotropic resolution. The normalized GM segments were then modulated for both linear and non-linear transformations and used for further analyses.

### Voxelwise and parcelwise MRI analysis

PD progression subtypes were compared using GMV in voxelwise and parcelwise space. Therefore, we used a linear model with the voxelwise or parcelwise GMV as the dependent variable and the subtype as the independent variable, integrating age, sex, total intracranial volume, and latent disease time as covariates. Visualization was done by coloring voxels/parcels based on their t-value when the difference between progression subtypes was significant. Visualizations were created based on test results prior to multiple testing correction, as indicated in the figure legends. Additionally, test results after correcting for multiple testing using Benjamini-Hochberg method are reported.^48^ For voxelwise GMV comparison, analysis was restricted to voxels inside the CAT 12.8 brain mask.

In a second approach, we applied additional 4-voxel smoothing using a full width at half maximum Gaussian kernel and 4-voxel resampling via linear interpolation to reduce the number of voxels and improve signal-to-noise ratio.

For parcelwise GMV comparison, we combined variants of Schaefer’s cortical atlas (100, 400, 800, or 1200 parcels)^49^, Fan’s subcortical atlas (36 parcels)^50^, and Buckner’s cerebellar atlas (37 parcels)^51^. We calculated the mean GMV of all voxels within each parcel (173, 473, 873, and 1273 total parcels).

Parcelwise and voxelwise analyses were executed using Python packages nibabel 5.2.1^52^ and nilearn 0.10.4^53^.

### Brain age models

Brain age models were selected from a recent publication that systematically compared 128 different brain age estimation workflows that combined different feature representations and machine learning algorithms.^26^ All brain age models were trained on the four HC datasets described above.

We selected three models from this publication, each performing best on a specific task:

1. Within-dataset brain age estimation (lowest mean absolute error): voxelwise feature representation using 4 mm smoothing and 4 mm resampling with Gaussian process regression (S4_R4+GPR)
2. Cross-dataset brain age estimation (lowest mean absolute error): voxelwise feature representation using 4 mm smoothing, 4 mm resampling, and subsequent principal component analysis with Gaussian process regression (S4_R4+PCA+GPR)
3. Correlation with clinical scores (highest Pearson’s correlation): voxelwise feature representation using no smoothing and 4 mm resampling with lasso regression (S0_R4+LR)

Several studies have shown that brain age is often overestimated in elderly individuals and underestimated in younger individuals.^54^ To remove this age-related bias, we compared two bias correction methods applied to the brain age estimations:

1. Cole et al. method^55^: linear model that estimates bias correction using estimated brain age
2. Beheshti et al. method^27^: linear model that estimates bias correction using BAG and chronological age

To select the optimal brain age estimation workflow for our PPMI dataset, we applied all three brain age models to estimate brain age on the PPMI HC cohort. Subsequently, brain age was corrected using the two bias correction methods described above in a 5-fold cross-validation fashion. We selected the workflow with the lowest mean absolute error on the HC test sets. For subsequent use in the PD cohort, we retrained the best bias correction method on the entire PPMI HC cohort.

The final selected brain age estimation workflow was further validated for re-test reliability and longitudinal consistency. We calculated the concordance correlation coefficient for BAG, assuming that BAG should remain relatively constant over short intervals. Therefore, we used longitudinal MRI scans with intervals of less than 1.5 years. Longitudinal consistency was assessed by comparing the increase in brain age between two subsequent MRI scans and the actual time between both MRI scans, using longitudinal MRI with more than 1.5 years in-between. If more than two MRI scans were available for an individual, the first and last MRI were used.

### BAG group comparisons

BAG differences between HC, the fast-progressing subtype, and the slow-progressing subtype were compared pairwise using a linear model with BAG as the dependent variable and group as the independent variable. Age and sex were included as covariates. For comparison of the fast-progressing and slow-progressing subtype, latent disease time was included additionally as a covariate.

BAG progression was analyzed by fitting a linear mixed-effects model for each group, with BAG as the dependent variable and time since first MRI as the independent variable. Fixed and patient-specific random intercepts and slopes were modeled.

### BAG correlations with clinical scores

BAG was correlated with (1) baseline outcomes and (2) progression of outcomes. For both analyses, some clinical scores were inverted so that higher scores always indicated more severe impairment or faster progression. Partial correlation coefficients were calculated with age as a covariate, using the Python package pingouin 0.5.4.^56^ For correlations with baseline outcomes, latent disease time was included as an additional covariate. Progression of outcomes was modeled on the latent disease timescale by fitting a linear mixed-effects model. Fixed and patient-specific random intercepts and slopes were included.

Correlation coefficients and respective p-values were calculated for the following outcomes: (1) clinical scores, (2) DaTSCAN uptake ratios, and (3) cerebrospinal fluid parameters. Benjamini-Hochberg correction for multiple testing was performed within each outcome group.^48^ DaTSCAN uptake ratios were analyzed separately for caudate nucleus, putamen, and striatum with (1) mean of left and right side and (2) asymmetry index as parameters. Cerebrospinal fluid parameters included amyloid beta 1-42, neurofilament light chain, phospho-tau, and phospho-tau / amyloid beta 1-42 ratio.

### Event-free survival analysis

Event-free survival analysis was performed using a Cox proportional hazards model for the events (1) occurrence of mild cognitive impairment (MCI), and (2) cognitive decline indicated by progression on an ordinal scale that included the items normal cognition, subjective cognitive impairment, mild cognitive impairment, and dementia. Modeling was performed using age at baseline visit, sex, and BAG at baseline as covariates with time modeled on the latent disease timescale. The proportional hazards assumption was checked by (1) performing statistical tests for any time-varying coefficients with p-values being corrected for multiple testing using the Benjamini-Hochberg method, and (2) visual inspection of scaled Schoenfeld residuals. The analysis was implemented using the Python package lifelines 0.30.0.^57^

### Sample size calculation

We assessed the effect of enriching people with PD with high baseline BAG in clinical cohorts by simulating an RCT with a disease-modifying treatment. Single cognitive scores or a cognitive compound score were used as the primary outcome. The cognitive compound score was calculated by (1) computing z-scores for cognitive outcomes based on the baseline mean and baseline standard deviation, (2) inverting z-scores for some outcomes so that higher scores always indicate more severe cognitive impairment, and (3) averaging the z-scores across cognitive tests. Therefore, we selected the four cognitive tests with the lowest sample size in the scenario without BAG stratification that were available at baseline (i.e., Hopkins Verbal Learning Test-Intermediate Recall, Letter Number Sequencing, Montreal Cognitive Assessment, Symbol Digit Modalities). Measurements were assumed every six months over two years. A power of 80% and a significance level of 0.1 were chosen. We assumed a treatment effect corresponding to a 30% reduction in the rate of disease progression (as measured by the cognitive composite score). For simplicity, we used equally sized control and treatment groups, excluding different treatment dosage arms. Power and sample size were calculated using the PPMI PD cohort and a linear mixed-effects model, following Edland et al. ^58^, implemented in R 4.3.2 with the longpower package 1.0.24.^59^

### Statistical analyses

All statistical tests were performed two-sided with a significance level of 0.05 using Python 3.12 with packages statsmodels 0.14.2^60^ and scikit-learn 1.5.0^61^. Boxplots display the median, interquartile range (IQR), whiskers extending to 1.5 times the IQR, and outliers as circles beyond the whiskers.

## Data availability

Clinical data is available from the PPMI study group (www.ppmi-info.org).

## Code availability

Source code for statistical analyses will be published at https://github.com/t-haehnel/Brainage-PD-Progression under the MIT license upon manuscript acceptance. Source code for brain age estimation workflows is available at https://github.com/juaml/brainage_estimation.

## Acknowledgments

No funding was obtained for conducting this study.

## Author contributions

T.H.: conceptualization, methodology, formal analysis, writing – original draft, visualization. F.H.: methodology, software, writing – review & editing. B.H.: supervision, writing – review & editing, H.F.: supervision, writing – review & editing, K.R.: writing – review & editing. S.M.: methodology, writing – review & editing

## Supplementary material

Supplementary material providing additional plots and method details is available online.

