## Supplemental file for "Brain Age Gap as Predictor of Disease Progression in Parkinson’s Disease"

### Supplementary Materials

Tom Hähnel[1,2], Shammi More[2], Felix Hoffstaedter[3,4], Kaustubh R. Patil[3,4], Holger Fröhlich[2,5,6]*, Björn Falkenburger[1,7]*

1. Department of Neurology, Medical Faculty and University Hospital Carl Gustav Carus, TUD Dresden University of Technology, Dresden, Germany
2. Department of Bioinformatics, Fraunhofer Institute for Algorithms and Scientific Computing (SCAI), Sankt Augustin, Germany
3. Institute of Neuroscience and Medicine (INM-7), Research Centre Jülich, Jülich, Germany
4. Institute of Systems Neuroscience, Medical Faculty, Heinrich Heine University Düsseldorf, Düsseldorf, German
5. Bonn-Aachen International Center for IT, University of Bonn, Bonn, Germany
6. Institute for Digital Medicine, University Medical Center Bonn, Bonn, Germany
7. German Center for Neurodegenerative Diseases (DZNE), Dresden, German

* Both authors contributed equally.

<u>Corresponding author:</u>
Dr. med. Tom Hähnel
Department of Neurology
Medical Faculty and University Hospital Carl Gustav Carus
TUD Dresden University of Technology
Fetscherstrasse 74
01307, Dresden, Germany


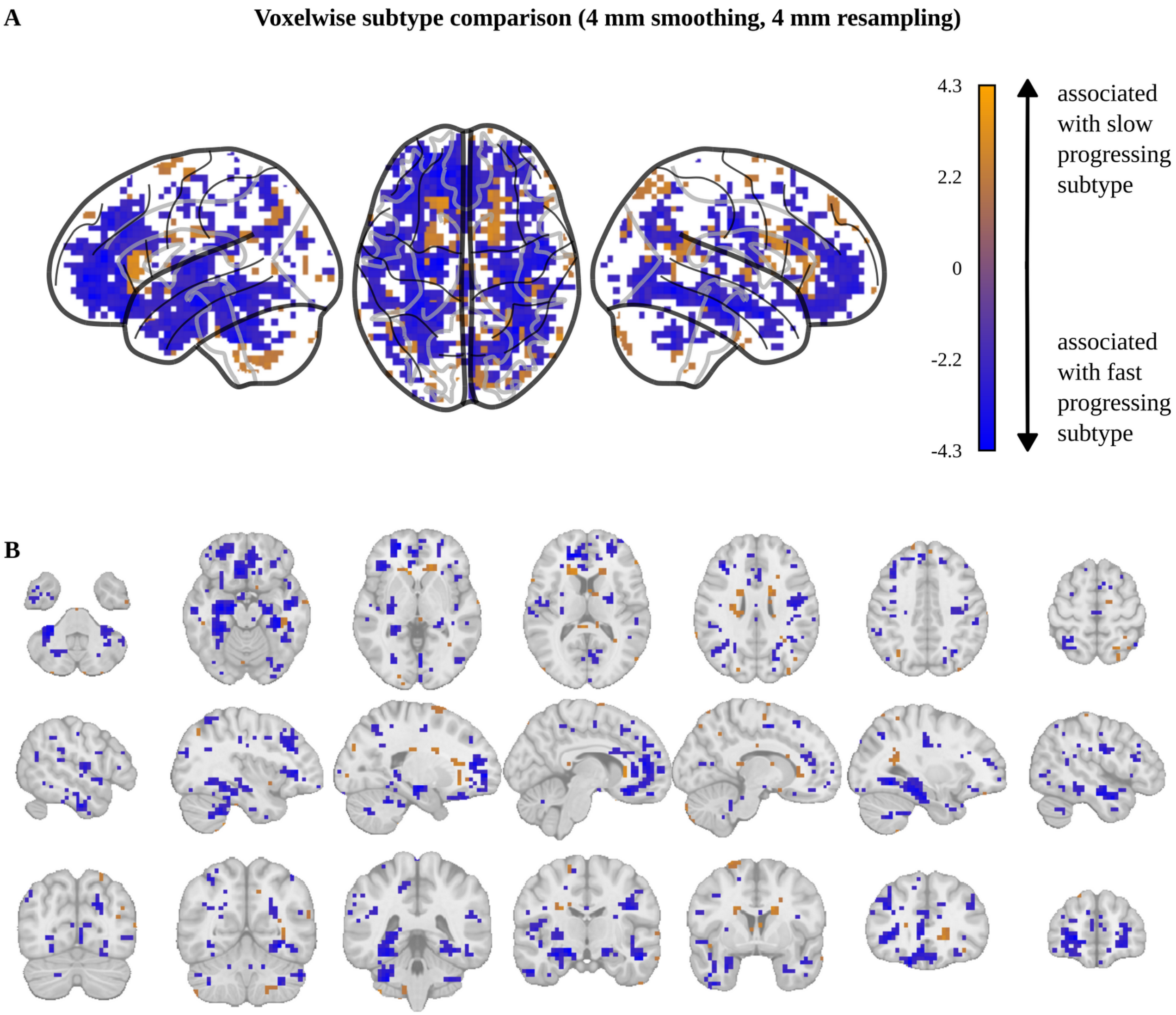


***Figure S1: Voxelwise gray matter volume comparison of PD progression subtypes using 4 mm smoothing and 4 mm resampling***

*Association of atrophy with the fast-progressing (blue) and slow-progressing (orange) subtypes displayed as* ***A)*** *glass brain visualization and* ***B)*** *multiple sagittal, coronal, and transverse planes. Voxels with significant progression subtype differences before correction for multiple testing are colored depending on their t-value (blue: associated with fast-progressing subtype, orange: associated with slow-progressing subtype). Note that no voxels remained significant after Benjamini-Hochberg correction for multiple testing. Additional 4 mm smoothing and 4 mm resampling were performed to reduce the number of features and to increase the signal-to-noise ratio.*

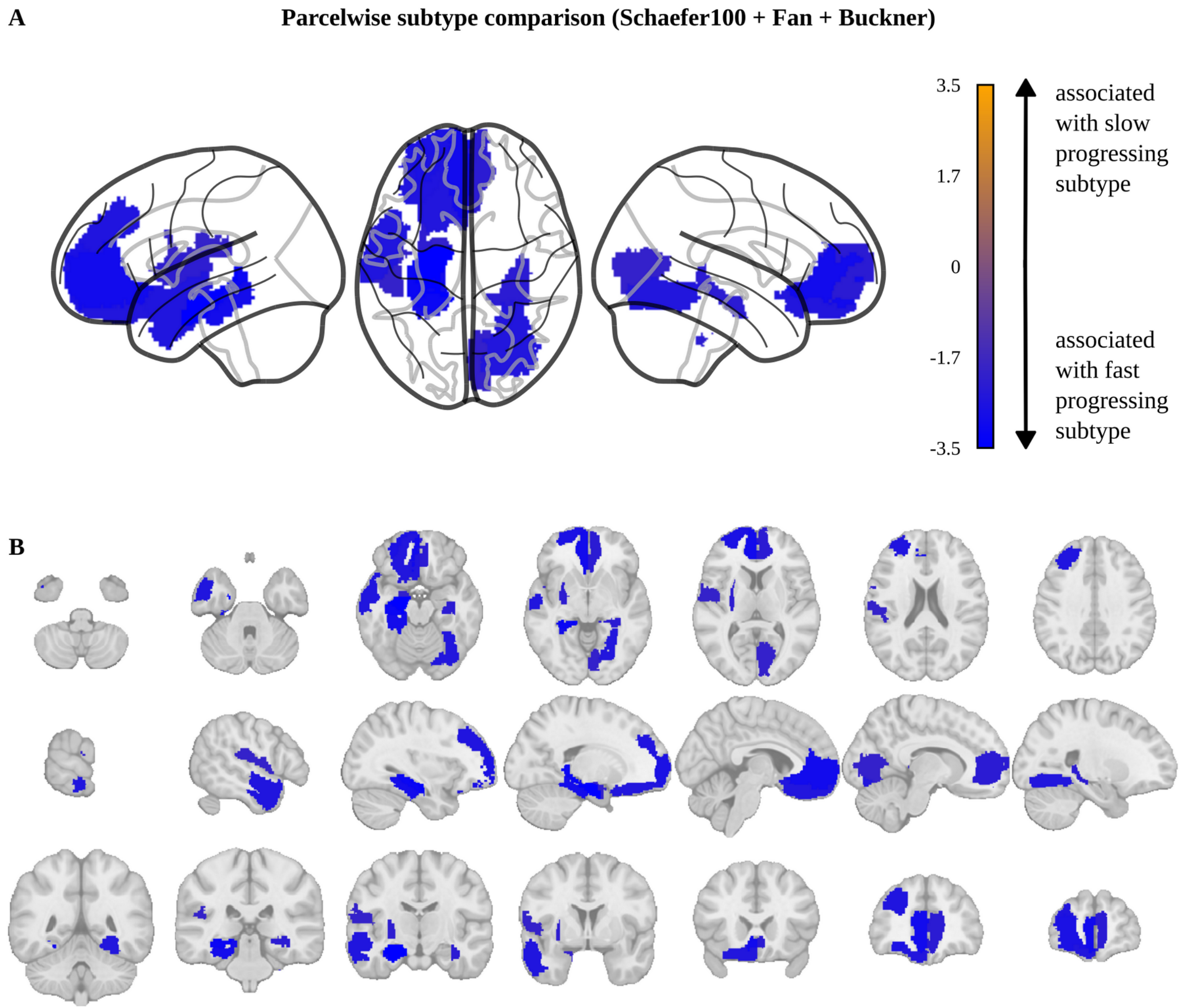


***Figure S2: Parcelwise gray matter volume comparison of PD progression subtypes using Schaefer100, Fan, and Buckner atlases***

*Association of atrophy with the fast-progressing (blue) and slow-progressing (orange) subtypes displayed as* ***A)*** *glass brain visualization and* ***B)*** *multiple sagittal, coronal, and transverse planes. Parcels with significant progression subtype differences before correction for multiple testing are colored depending on their t-value (blue: associated with fast-progressing subtype, orange: associated with slow-progressing subtype). Note that no parcels remained significant after Benjamini-Hochberg correction for multiple testing. A combination of Schaefers atlas with 100 cortical parcels, Fan atlas with 36 subcortical parcels, and Buckner atlas with 37 cerebellar parcels has been used, resulting in 173 parcels overall.*

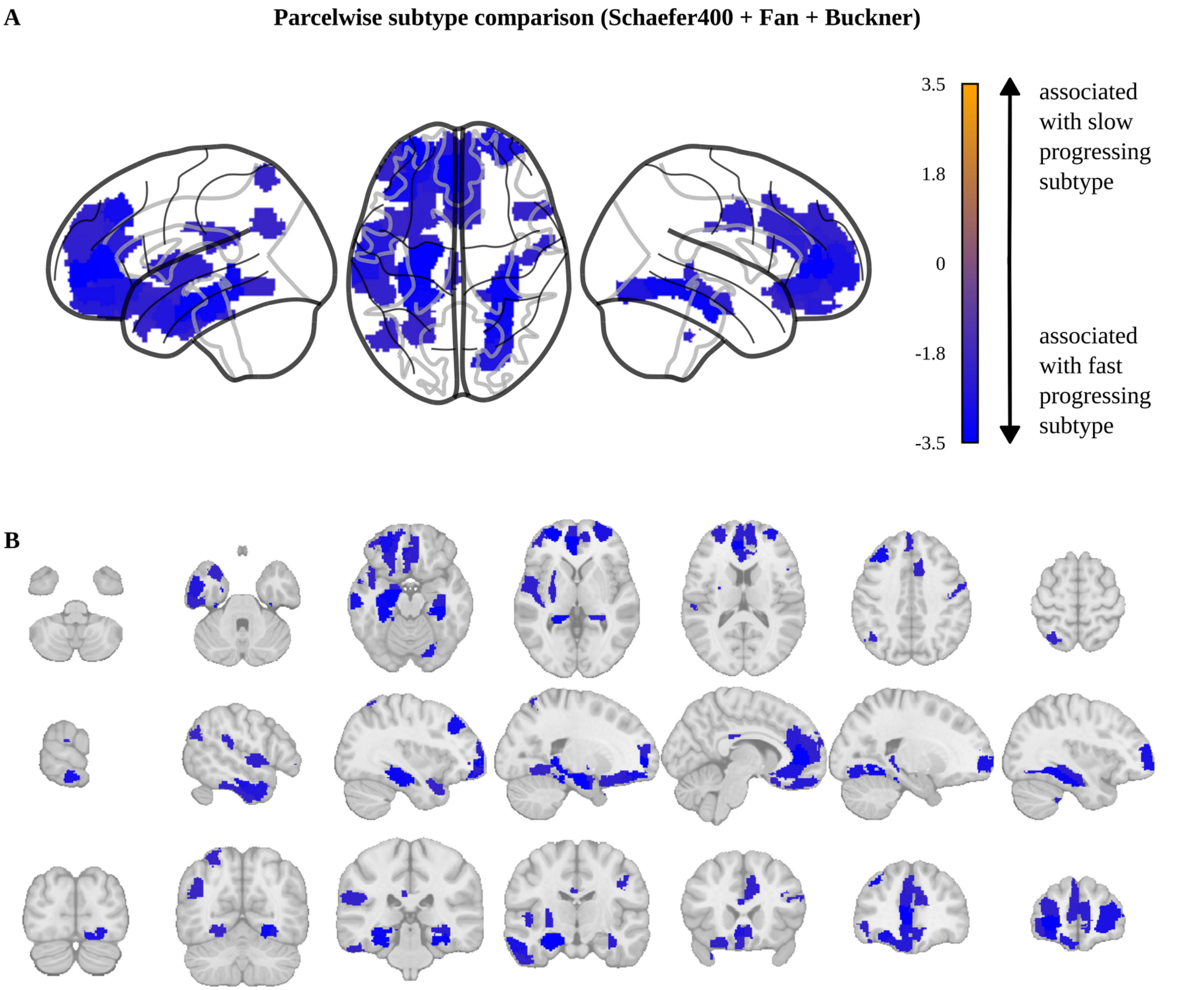


***Figure S3: Parcelwise gray matter volume comparison of PD progression subtypes using Schaefer400, Fan, and Buckner atlases***

*Association of atrophy with the fast-progressing (blue) and slow-progressing (orange) subtypes displayed as **A)** glass brain visualization and **B)** multiple sagittal, coronal, and transverse planes. Parcels with significant progression subtype differences before correction for multiple testing are colored depending on their t-value (blue: associated with fast-progressing subtype, orange: associated with slow-progressing subtype). Note that no parcels remained significant after Benjamini-Hochberg correction for multiple testing. A combination of Schaefers atlas with 400 cortical parcels, Fan atlas with 36 subcortical parcels, and Buckner atlas with 37 cerebellar parcels has been used, resulting in 473 parcels overall.*

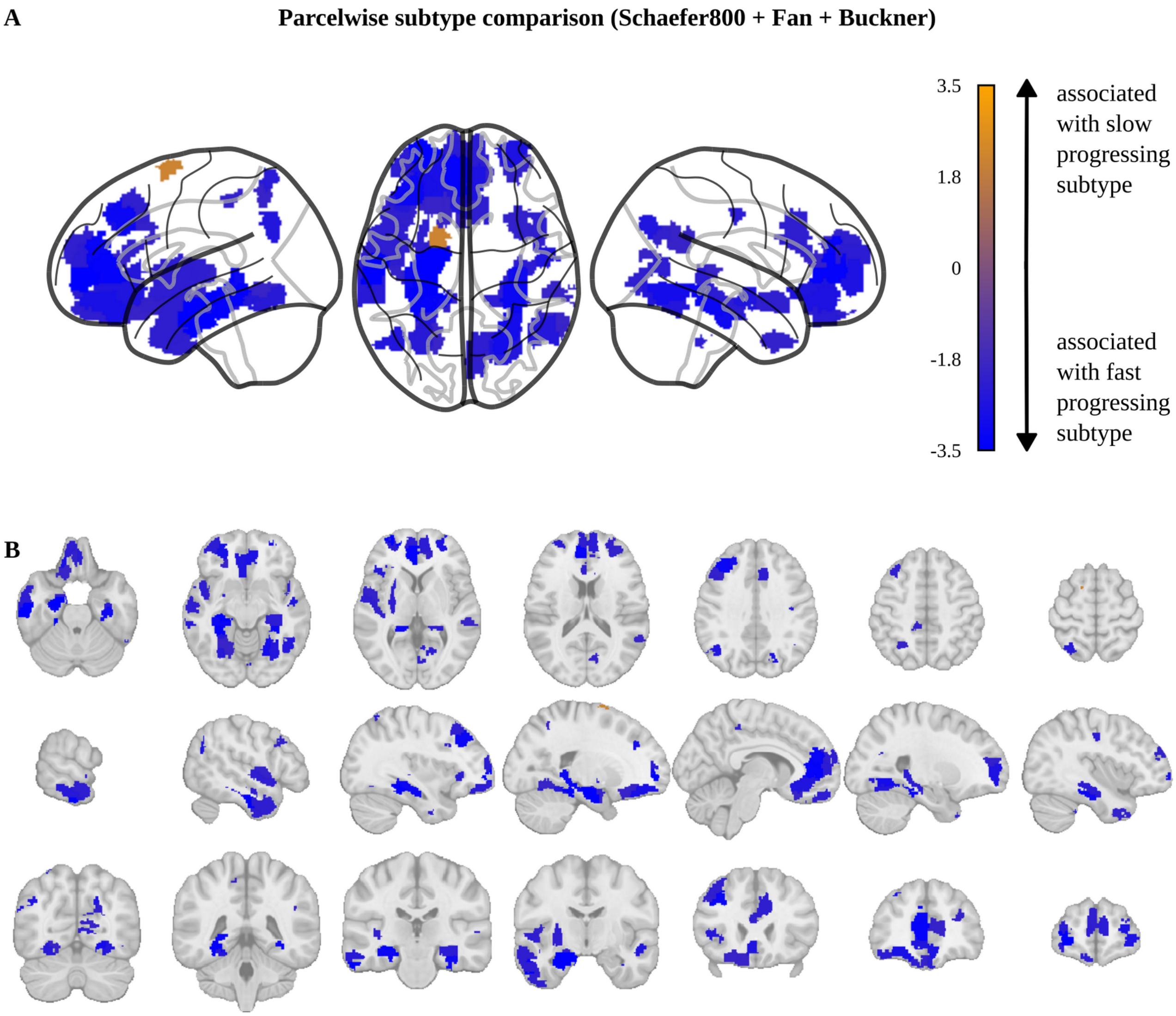


***Figure S4: Parcelwise gray matter volume comparison of PD progression subtypes using Schaefer800, Fan, and Buckner atlases***

*Association of atrophy with the fast-progressing (blue) and slow-progressing (orange) subtypes displayed as* ***A)*** *glass brain visualization and* ***B)*** *multiple sagittal, coronal, and transverse planes. Parcels with significant progression subtype differences before correction for multiple testing are colored depending on their t-value (blue: associated with fast-progressing subtype, orange: associated with slow-progressing subtype). Note that no parcels remained significant after Benjamini-Hochberg correction for multiple testing. A combination of Schaefers atlas with 800 cortical parcels, Fan atlas with 36 subcortical parcels, and Buckner atlas with 37 cerebellar parcels has been used, resulting in 873 parcels overall.*

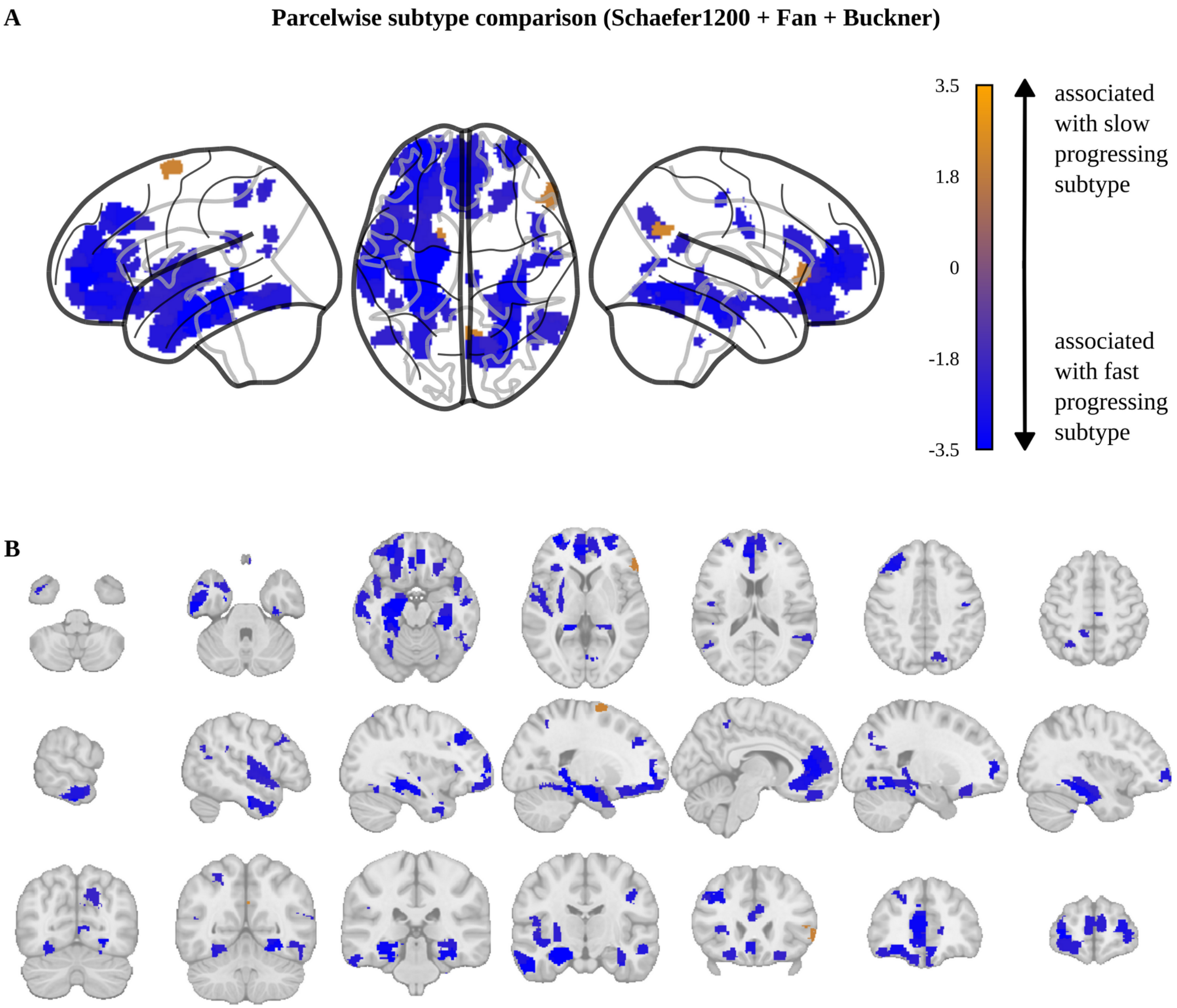


***Figure S5: Parcelwise gray matter volume comparison of PD progression subtypes using Schaefer1200, Fan, and Buckner atlases***

*Association of atrophy with the fast-progressing (blue) and slow-progressing (orange) subtypes displayed as **A)** glass brain visualization and **B)** multiple sagittal, coronal, and transverse planes. Parcels with significant progression subtype differences before correction for multiple testing are colored depending on their t-value (blue: associated with fast-progressing subtype, orange: associated with slow-progressing subtype). Note that no parcels remained significant after Benjamini-Hochberg correction for multiple testing. A combination of Schaefers atlas with 1200 cortical parcels, Fan atlas with 36 subcortical parcels, and Buckner atlas with 37 cerebellar parcels has been used, resulting in 1273 parcels overall.*

| Feature representation and machine learning model | Bias correction method | Mean absolute error (years) | Correlation brain age vs chronological age | Correlation BAG vs chronological age | Mean BAG (years) |
|---|---|---|---|---|---|
| S0_R4+LR | Beheshti | 4.51 | ρ=0.89 (p<0.0001) | p=0.95 | -0.01 |
| S4_R4+GPR | Beheshti | 4.71 | ρ=0.89 (p<0.0001) | p=0.95 | -0.01 |
| S4_R4+PCA+GPR | Beheshti | 5.01 | ρ=0.87 (p<0.0001) | p=0.95 | -0.01 |
| S4_R4+GRP | Cole | 5.06 | ρ=0.87 (p<0.0001) | p=1.0 | 0.0 |
| S4_R4+PCA+GPR | Cole | 5.08 | ρ=0.87 (p<0.0001) | p=1.0 | 0.0 |
| S0_R4+LR | Cole | 5.17 | ρ=0.86 (p<0.0001) | p=1.0 | 0.0 |

***Table S1: Brain age workflow selection on PPMI HC***
*To select the optimal brain age estimation workflow for our PPMI dataset, we applied three brain age models to estimate brain age on the PPMI HC cohort, comparing different feature representations (S0: no smoothing, S4: 4 mm smoothing, R4: 4 mm resampling, PCA: additional principal component analysis) and machine learning methods (GPR: Gaussian process regression, LR: lasso regression). Subsequently, brain age was corrected using two bias correction methods (Beheshti, Cole) in a 5-fold cross-validation fashion. We selected the workflow with the lowest mean absolute error on the HC test sets. In addition, the following statistics are shown: Pearson correlation for brain age vs. chronological age; p-values for Pearson correlation of BAG with chronological age; and mean BAG.*
*Abbreviations: BAG: brain age gap.*

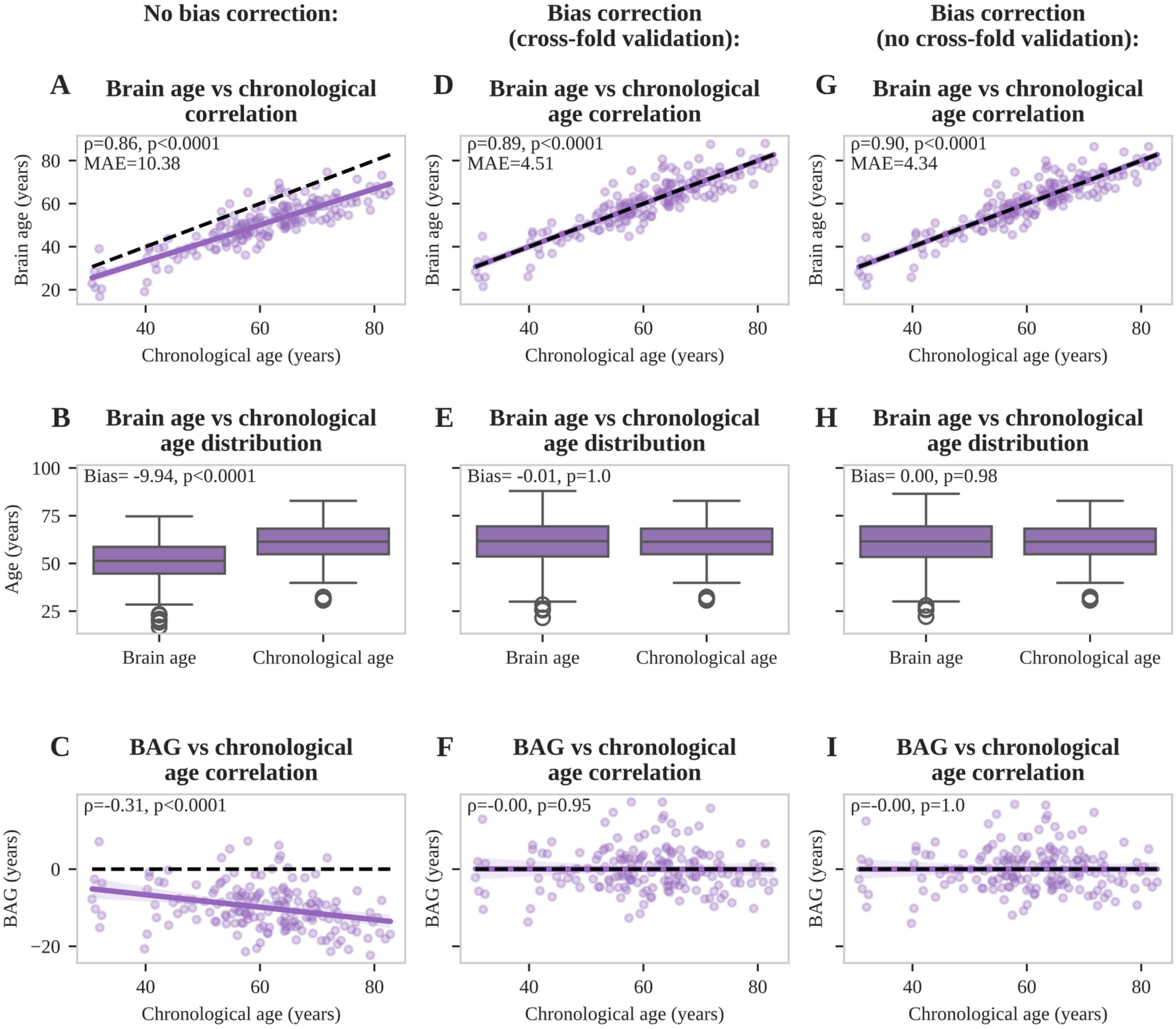


***Figure S6: Bias correction and validation on PPMI HC***

*Bias correction process for the finally selected brain age model (S0_R4+LR) using the Beheshti method and PPMI HC data. Without bias correction, brain age estimations are highly correlated with chronological age* ***(A)****, but show a bias (****B****) towards higher BAG in elderly people (****C****). Model selection was performed using bias correction on the PPMI HC data in a cross-validation framework with evaluation on test sets (****D****). The cross-validation approach confirmed a complete bias removal (****E****) with no residual correlation between BAG and chronological age in the test sets (****F****). Finally, the bias correction model was retrained on the whole PPMI HC dataset (****G-I****), before applying it to the PPMI PD cohort.*

*Abbreviations: BAG: brain age gap.*

| Clinical score | Score at baseline | Score progression |
|---|---|---|
| Boston Naming Test | .. | -0.063 (p=0.59) |
| Hopkins Verbal Learning Test DR | 0.023 (p=0.73) | **-0.137 (p=0.044)** |
| Hopkins Verbal Learning Test IR | **-0.134 (p=0.036)** | **-0.150 (p=0.03)** |
| Judgement Line Orientation | **-0.154 (p=0.028)** | -0.076 (p=0.39) |
| Letter Number Sequencing | **-0.132 (p=0.036)** | **-0.178 (p=0.019)** |
| MoCA | **-0.135 (p=0.036)** | **-0.135 (p=0.044)** |
| Symbol Digit Modalities | -0.028 (p=0.73) | **-0.154 (p=0.03)** |
| Trailmaking A | .. | 0.081 (p=0.47) |
| Trailmaking B | .. | 0.032 (p=0.83) |
| VFT phonematic F | 0.048 (p=0.58) | **-0.158 (p=0.03)** |
| VFT semantic animal | -0.061 (p=0.5) | -0.124 (p=0.067) |
| VFT semantic fruits | -0.037 (p=0.73) | **-0.145 (p=0.034)** |
| VFT semantic vegetables | -0.065 (p=0.48) | -0.099 (p=0.2) |
| CGI | .. | 0.056 (p=0.83) |
| PGI | .. | -0.124 (p=0.52) |
| SEADL | **-0.186 (p=0.0047)** | -0.034 (p=0.75) |
| MDS-UPDRS III axial off | **0.133 (p=0.036)** | 0.018 (p=0.87) |
| H&Y off | 0.053 (p=0.55) | -0.057 (p=0.5) |
| PIGD off | 0.005 (p=0.92) | 0.038 (p=0.71) |
| MDS-UPDRS II | -0.020 (p=0.73) | 0.060 (p=0.5) |
| MDS-UPDRS III off | **0.189 (p=0.0047)** | -0.007 (p=0.93) |
| ESS | -0.021 (p=0.73) | 0.024 (p=0.83) |

| | | |
|---|---|---|
| RBD-SQ | 0.082 (p=0.29) | 0.067 (p=0.45) |
| SCOPA-AUT | 0.023 (p=0.73) | -0.000 (p=0.99) |
| MDS-UPDRS I | -0.034 (p=0.73) | 0.091 (p=0.25) |
| MDS-UPDRS I-III off | **0.143 (p=0.036)** | 0.017 (p=0.87) |
| GDS | 0.020 (p=0.73) | -0.011 (p=0.93) |
| QUIP | -0.120 (p=0.061) | -0.007 (p=0.93) |
| STA | 0.056 (p=0.53) | 0.074 (p=0.39) |

***Table S2: Correlation of BAG with baseline clinical scores and progression of clinical scores***
*Correlation coefficients and corresponding p-values are reported for partial correlations of clinical scores with BAG in the PD group. Correlations were calculated for baseline values (left column) and progression of scores (right column). Correlation coefficients were corrected for age (baseline and progression) and latent disease time (baseline). Corresponding p-values were corrected for multiple testing using the Benjamini-Hochberg method. Two dots (..) indicate that the corresponding score was not measured at baseline. Significant results are indicated in bold.*
*Abbreviations: BAG: brain age gap, CGI: clinical global impression scale, DR: delayed recall, ESS: Epworth Sleepiness Scale, GDS: Geriatric Depression Scale, H&Y: Hoehn & Yahr, IR: intermediate recall, MDS-UPDRS: MDS-Unified Parkinson's Disease Rating Scale, MoCA: Montreal Cognitive Assessment, PGI: Patient Global Impression scale, PIGD: Postural Instability/Gait Disturbance, QUIP: Questionnaire for Impulsive-Compulsive Disorders in Parkinson's Disease Rating Scale, RBD-SQ: REM Sleep Behavior Disorder-Screening Questionnaire, SCOPA: Scales for Outcomes in Parkinson's disease - Autonomic dysfunction, SEADL: Schwab and England Activities of Daily Living scale, STA: State-Trait Anxiety Inventory, VFT: Verbal Fluency Task.*

| DaTSCAN parameter | Parameter at baseline | Parameter progression |
|---|---|---|
| Ncl. Caudatus | 0.012 (p=0.92) | **-0.144 (p=0.036)** |
| Ncl. Caudatus asymmetry | -0.045 (p=0.81) | -0.008 (p=0.88) |
| Putamen | -0.008 (p=0.92) | -0.067 (p=0.31) |
| Putamen asymmetry | -0.074 (p=0.76) | 0.080 (p=0.26) |
| Striatum | 0.005 (p=0.92) | **-0.133 (p=0.036)** |
| Striatum asymmetry | -0.062 (p=0.76) | 0.024 (p=0.78) |

***Table S3: Correlation of BAG with DaTSCAN parameters***

*Correlation coefficients and corresponding p-values are reported for partial correlations of DaTSCAN uptake ratios with BAG in the PD group. Correlations were calculated for baseline parameters (left column) and progression of DaTSCAN parameters (right column). Correlation coefficients were corrected for age (baseline and progression) and latent disease time (baseline). Corresponding p-values were corrected for multiple testing using the Benjamini-Hochberg method. Significant results are indicated in bold.*

*Abbreviations: BAG: brain age gap.*

| Cerebrospinal fluid parameter | Parameter at baseline | Parameter progression |
|---|---|---|
| A Beta 1-42 | -0.083<br>(p=0.49) | 0.068<br>(p=0.31) |
| Neurofilament light chain | 0.041<br>(p=0.61) | 0.117<br>(p=0.31) |
| pTau | -0.028<br>(p=0.61) | 0.008<br>(p=0.88) |
| pTau / A Beta 1-42 | 0.061<br>(p=0.55) | 0.066<br>(p=0.31) |

***Table S4: Correlation of BAG with cerebrospinal fluid parameters***

*Correlation coefficients and corresponding p-values are reported for partial correlations of cerebrospinal fluid parameters with BAG in the PD group. Correlations were calculated for baseline (left column) and progression (right column) of cerebrospinal fluid parameters. Correlation coefficients were corrected for age (baseline and progression) and latent disease time (baseline). Corresponding p-values were corrected for multiple testing using the Benjamini-Hochberg method. Significant results are indicated in bold.*

*Abbreviations: BAG: brain age gap.*

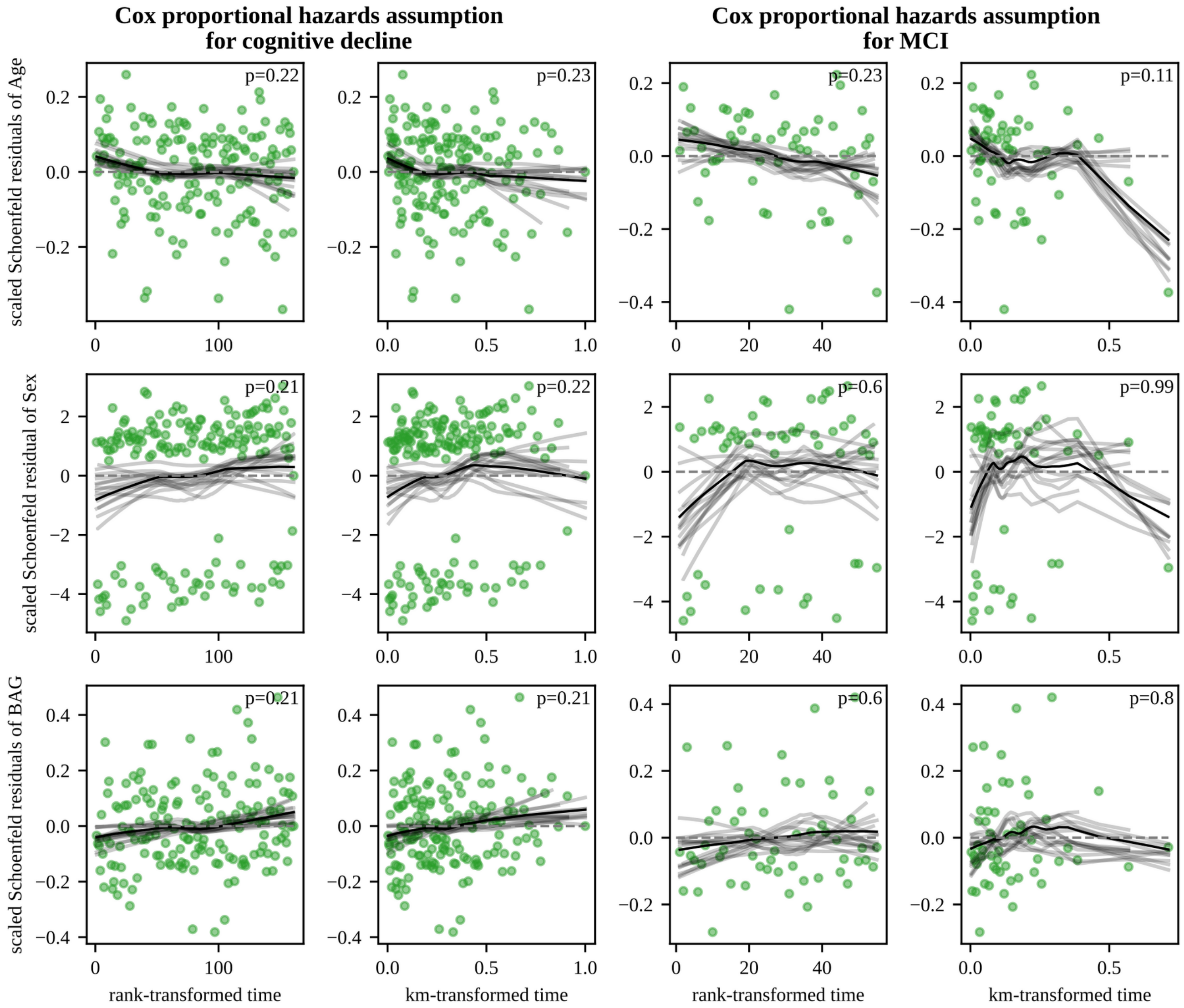


***Figure S7: Scaled Schoenfeld residual plots for checking the proportional hazards assumption***
*Scaled Schoenfeld residuals are presented for the Cox proportional hazards model of event-free survival analysis for both cognitive decline (left two columns) and mild cognitive impairment (right two columns). For each covariate (age, sex, BAG), plots and statistics are shown for rank-based and Kaplan-Meier based time transformations. P-values to test for any time-varying coefficients are reported and were corrected for multiple testing using Benjamini-Hochberg method. Visualizations are based on the Python lifelines package. Residuals are presented as green dots. The black line was derived using locally weighted smoothing (lowess), with gray lines derived via additional bootstrapping.*
*Abbreviations: BAG: brain age gap, km: Kaplan-Meier.*

| Clinical score | Sample size (no stratification) | Sample size reduction (50th percentile) | Sample size reduction (70th percentile) | Sample size reduction (90th percentile) |
|---|---|---|---|---|
| Letter Number Sequencing | 5327 | 36 % | 42 % | 75 % |
| VFT phonematic | 888894 | 88 % | 96 % | 99 % |
| VFT semantic animal | 7811 | 42 % | 57 % | 44 % |
| VFT semantic fruits | 246805 | 93 % | 96 % | 97 % |
| VFT semantic vegetables | 13453 | 52 % | 64 % | 75 % |
| Hopkins Verbal Learning Test DR | 133867 | 75 % | 81 % | 93 % |
| Hopkins Verbal Learning Test IR | 4027 | 13 % | 18 % | 48 % |
| MoCA | 4937 | 47 % | 40 % | 61 % |
| Judgment Line Orientation | 28561 | 42 % | 52 % | 50 % |
| Symbol Digit Modalities | 1724 | 34 % | 31 % | 66 % |
| Cognitive composite score | 1212 | 23 % | 28 % | 58 % |

***Table S5: Sample size reductions for different cognitive primary outcomes***
*Sample size without baseline stratification and relative sample size reductions for patient stratification based on 50th percentile, 70th percentile, and 90th percentile BAG at baseline.*
*Abbreviations: BAG: brain age gap, DR: delayed recall, IR: intermediate recall, MoCA: Montreal Cognitive Assessment, VFT: Verbal Fluency Task.*

| Feature representation and machine learning model | Bias correction method | Mean BAG (years) |
| --- | --- | --- |
| S0_R4+LR | Beheshti | 1.06 (p=0.00062) |
| S4_R4+GPR | Beheshti | 1.06 (p=0.00012) |
| S4_R4+PCA+GPR | Beheshti | 1.14 (p=0.00012) |
| S0_R4+LR | Cole | 1.26 (p=0.00062) |
| S4_R4+GPR | Cole | 1.18 (p=0.00012) |
| S4_R4+PCA+GPR | Cole | 1.19 (p=0.00012) |

***Table S6: BAG for the PPMI PD cohort using different brain age estimation workflows***
*Comparison of mean BAG for the PPMI PD cohort using the six workflows described in the manuscript. Bias correction methods were always trained on the whole HC group. Mean BAG with corresponding p-value from a pairwise t-test comparing chronological age and brain age are reported.*
*Abbreviations: BAG: brain age gap, GPR: Gaussian process regression, LR: lasso regression, PCA: principal component analysis, R4: 4 mm resampling, S0: no smoothing, S4: 4 mm smoothing.*
*Abbreviations: BAG: brain age gap.*
